# Impact of Case Detection and COVID-19-Related Disruptions on Tuberculosis in Vietnam: A Modelling Analysis

**DOI:** 10.1101/2025.02.27.25323053

**Authors:** Viet Long Bui, Romain Ragonnet, Angus E. Hughes, David S. Shipman, Emma S. McBryde, Binh Hoa Nguyen, Hoang Nam Do, Thai Son Ha, Greg J. Fox, James M. Trauer

**Author notes:** Corresponding author:* Viet Long Bui, School of Public Health and Preventive Medicine, Monash University, 553 St Kilda Road, Melbourne, VIC 3004, Australia.

## Abstract

**Background:** Vietnam, a high-burden tuberculosis (TB) country, experienced marked declines in TB notifications during the COVID-19 pandemic. We assessed the impact of pandemic-related disruptions on TB case detection and transmission using a dynamic transmission model calibrated to local demographic and epidemiological observations.

**Methods:** We developed an age-structured compartmental TB transmission model to estimate COVID-19’s impact on TB in Vietnam. Four model assumptions reflecting reductions in detection and/or transmission were calibrated to notification data, with the best-fitting assumption used for future projections and to evaluate the effects of enhanced case detection scenarios.

**Results:** COVID-19 significantly disrupted TB services in Viet Nam, resulting in an estimated 2,000 additional TB episodes (95% credible interval [CrI]: 200-5,100) and 1,100 TB-related deaths (95%CrI: 100-2,700) in 2021.By 2035, the cumulative impact of these disruptions could reach 22,000 additional TB episodes (95%CrI: 2,200-63,000) and 5,900 deaths (95%CrI: 600-16,600) by 2035. We predicted two hypothetical scenarios of enhancing TB case detection. Under the ambitious scenario, enhancing TB case detection could mitigate these potential impacts by preventing 17.8% of new TB episodes (95%CrI: 13.1%-21.9%) and 34.2% (95%CrI: 31.5%-37.0%) of TB-related deaths by 2035, compared to no enhancement.

**Conclusions:** COVID-19-related disruptions have hindered TB detection in Vietnam, likely causing long-term increases in new TB episodes and deaths. However, the uncertainty around these effects is considerable. Sustained investment in diagnostics, system resilience, and patient-centric policies have the potential to achieve benefits that are substantially larger than these pandemic-related setbacks.

**Summary:** COVID-19 disrupted TB detection in Vietnam, leading to long-term increases in incidence and mortality. Modelling suggests improved case-finding could avert a sizeable portion of new infections and deaths, but sustained investment is essential to improve TB prevention and care.

## INTRODUCTION

Tuberculosis (TB), the disease caused by the bacterium *Mycobacterium tuberculosis (M.tb)*, remains a critical global public health challenge [1]. Vietnam is among the 30 high TB burden countries designated by the World Health Organization (WHO), with an estimated incidence of 182 per 100,000 population in 2023 [1]. Despite decades of programmatic effort and sustained political commitment, TB control in Vietnam remains a challenge due to a combination of delayed case detection, underdiagnosis, and gaps in access to care.

The COVID-19 pandemic appears to have severely disrupted global TB prevention and care, causing significant declines in TB notifications in high-burden countries [2–4]. The WHO reported an 18% global decline in TB notifications in 2020, with an estimated 200,000–400,000 additional TB deaths [2, 5]. WHO TB burden estimates are primarily based on static models using notification trends, prevalence surveys, or expert adjustments, and only recently incorporated dynamic transmission models for a limited number of countries significantly affected by COVID-19 [6], which may not be the optimal approach to predicting long-term TB impact estimates. The reduction in TB notifications may be attributable to healthcare disruptions, resource diversion, reduced healthcare-seeking behavior or some combination of these factors. However, COVID-19 measures like mask-wearing and social distancing may also have reduced TB transmission, which are not accounted for in WHO’s early estimates [5]. Vietnam implemented several stringent public health and social measures (PHSMs) in the region, including aggressive contact tracing, lockdowns, and mobility restrictions, particularly during 2021 [7]. Given these complex and context-specific dynamics, a dynamic modelling approach is essential to accurately quantify the impact of COVID-19 on *M.tb* transmission and TB case detection in Vietnam.

In this study, we developed a dynamic TB transmission model calibrated to Vietnam’s demographic and epidemiological data to simulate *M.tb* transmission dynamics, estimate the impact of COVID-19 on *M.tb* transmission, and project the effects of hypothetical scenarios involving enhanced case finding.

## METHODS

### 1. Model structure

We developed a compartmental model comprising six compartment types - Susceptible (S), Early latent (E), Late latent (L), Active TB or infectious (I), Treatment (T), and recovered (R) - to represent different TB-related infection and disease states, using a similar conceptual approach and assumptions as our previously published model [8]. The model was stratified by age, BCG vaccination, and clinical forms of TB, categorized into three clinical severity groups, which we termed smear-positive TB (SPTB), smear-negative TB (SNTB), and extrapulmonary TB (EPTB). These groups are defined primarily by differences in natural history and infectiousness, with SPTB patients characterized by a more rapid disease progression and high infectiousness. By contrast, SNTB represents a more benign disease course with partial infectiousness, while EPTB is considered non-infectious. A detailed description of the model structure and all associated methods are provided in the Supplemental Material.

The reduction in TB notifications during the COVID-19 pandemic could plausibly be attributable to reduced case detection, decreased transmission, or both. To model these effects, we considered four candidate assumptions: Assumption 1, the COVID-19 pandemic had no impact on either TB detection or transmission; Assumption 2, only case detection was reduced, reflecting the decline in healthcare access, while TB transmission remained unaffected; Assumption 3, only TB transmission was reduced, with public health and social measures (PHSMs) limiting transmission, but with case detection continuing unaffected; Assumption 4, both case detection and TB transmission were reduced, incorporating the effects of both PHSMs and healthcare disruptions.

Table S2 (Supplementary Material) presents model parameters. One of the key parameters in our model was the rate of commencing treatment for active TB, which governs the transition of individuals from (I) to (T). This rate is the reciprocal of the average time from developing active TB to first presentation and treatment initiation under the National TB Program (NTP). The average time from developing active TB to first presentation for care and treatment initiation reflects delays attributable to the patient, provider, and health system factors, which can each contribute to delayed diagnoses. We used a smooth transition function that gradually increased with the historical improvements in the availability of TB diagnostics and treatments over the pre-pandemic period.

In our model, the impact of COVID-19 on *M.tb* transmission and case detection were incorporated through the use of reduction factors with minimally informative uniform priors, ranged from 0.01 to 0.90. These multipliers gradually declined from 2020 to 2021, before returning to baseline by 2022. Under Assumptions 2 and 4, reductions affected treatment initiation; under Assumptions 3 and 4, they applied to contact rate. In Assumption 4, transmission and detection reductions were separately parameterized and calibrated to reflect distinct effect of the disruption.

We calibrated our model using the best available empirical data specific to Vietnam, including the national population size [9], TB notifications and the proportion of pulmonary TB among these notifications [10], and the prevalence of bacteriologically-confirmed and of SPTB among adults from the second national TB prevalence survey [11, 12] (Table S3, Supplementary Materials). The combined modelled prevalence of SPTB and SNTB was calibrated to match the bacteriologically-confirmed prevalence reported in the survey. We compared our model’s independent estimates of TB incidence and mortality with WHO’s estimates to support the plausibility of our projections.

The best assumption regarding COVID-19 impacts on *M.tb* transmission and case detection was identified by comparing the expected log pointwise predictive density Watanabe-Akaike Information Criterion (EPLD-WAIC) values across four scenarios, with less negative values indicating better predictive performance [13].

### 3. Future projections

Using the best-performing assumption, which captured the sharp COVID-19-related decline in TB notifications, as the ‘*status-quo’* scenario, we projected new TB episodes and TB-related deaths from 2025 to 2035, and compared these outcomes with a counterfactual scenario in which COVID-19 had no impact on *M.tb* transmission and TB case detection. Additionally, we assessed the potential effects of enhanced case detection scenarios, simulating two- and five-fold increases in TB treatment initiation rates by 2035. For each scenario, we estimated the number of new TB episodes and TB-related deaths averted.

### 4. Software

We used *summer*, a Python-based epidemiological modelling framework [14], and performed model calibration using PyMC-based DEMetropolisZ Bayesian inference algorithm [15]. Our model development, optimization, and calibration followed a pipeline equivalent to that detailed in [16]. The code used for the analyses is accessible in a GitHub repository at: https://github.com/vlbui/tbdynamics.

**Figure 1.**
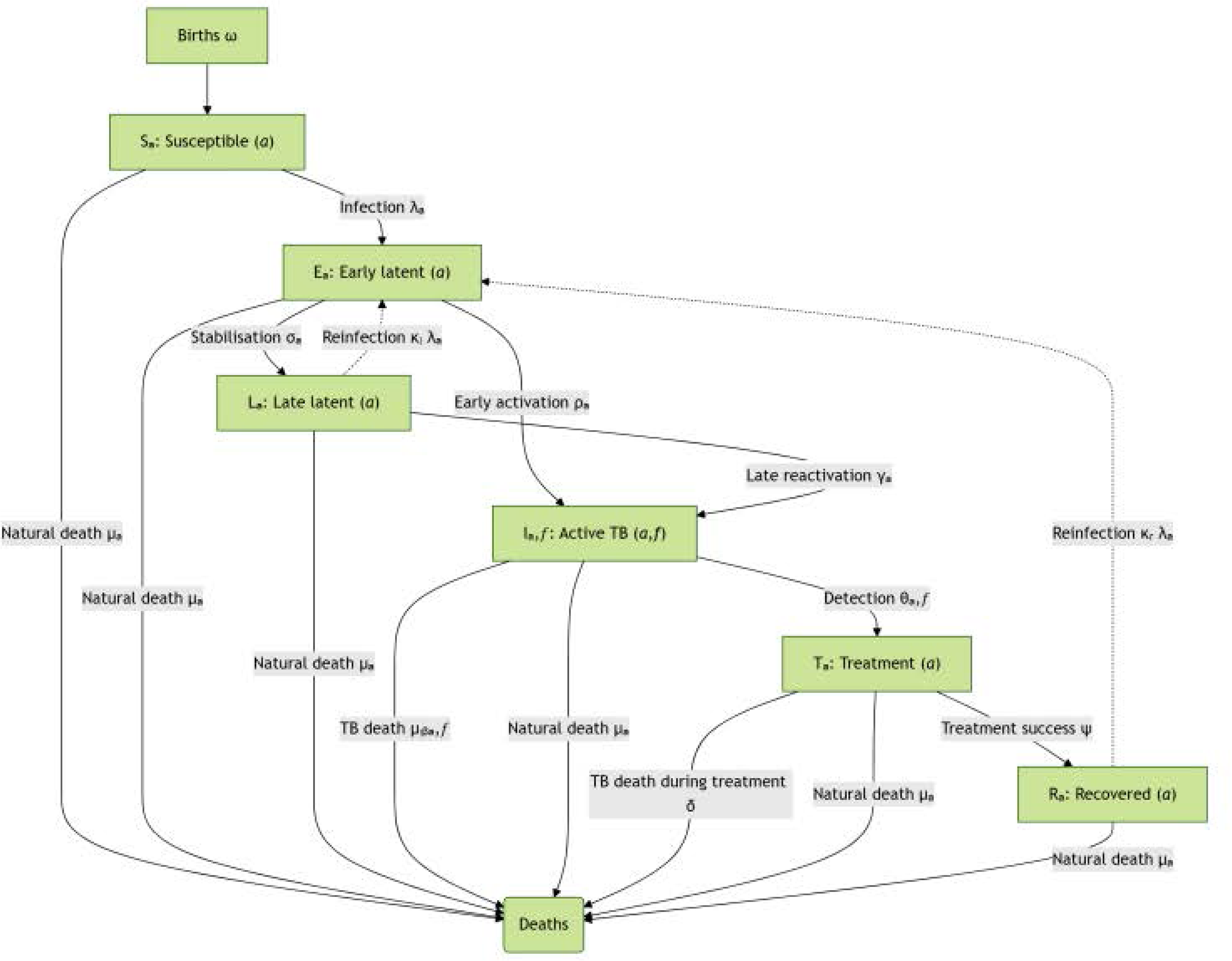
Model structure. Boxes represent different compartment types: Susceptible (S), Early latent infection (E), Late latent (L), Active TB (I), Treatment (T) and Recovered (R). Subscripts indicate stratification by age (a) and pulmonary/smear status (f). Alt text: The image depicts a compartmental model structure used in epidemiology. It consists of several boxes representing different disease states, including Susceptible (S), Early latent Infection (E), Late latent (L), Active TB (I), Treatment (T), and Recovered (R). Each box is further stratified by age (indicated by subscript ‘a’) and pulmonary/smear status (indicated by subscript ‘f’), illustrating the transitions and interactions between these different states in the model.

## RESULTS

### 1. Calibration results

#### Comparison of candidate assumptions

All four assumptions closely matched the total population and the prevalence of TB among adults, as shown in Figures S3 and S4 (Supplementary Material). However, the fit varied when comparing the actual number of TB notifications to the equivalent model output. Figure 2 shows this comparison under our four candidate assumptions regarding the effect of COVID-19 on *M.tb* dynamics. Among the candidate assumptions, Assumption 2, which assumed reduced TB detection during the COVID-19 pandemic, best fit the sharp decline in TB notifications in 2021, slightly outperforming Assumption 4 (reductions in both detection and transmission) based on EPLD-WAIC (Table S4, Supplementary Material). In contrast, Assumption 1 (no COVID-19 impact) and Assumption 3 (reduced transmission only) fit the data less well.

**Figure 2.**
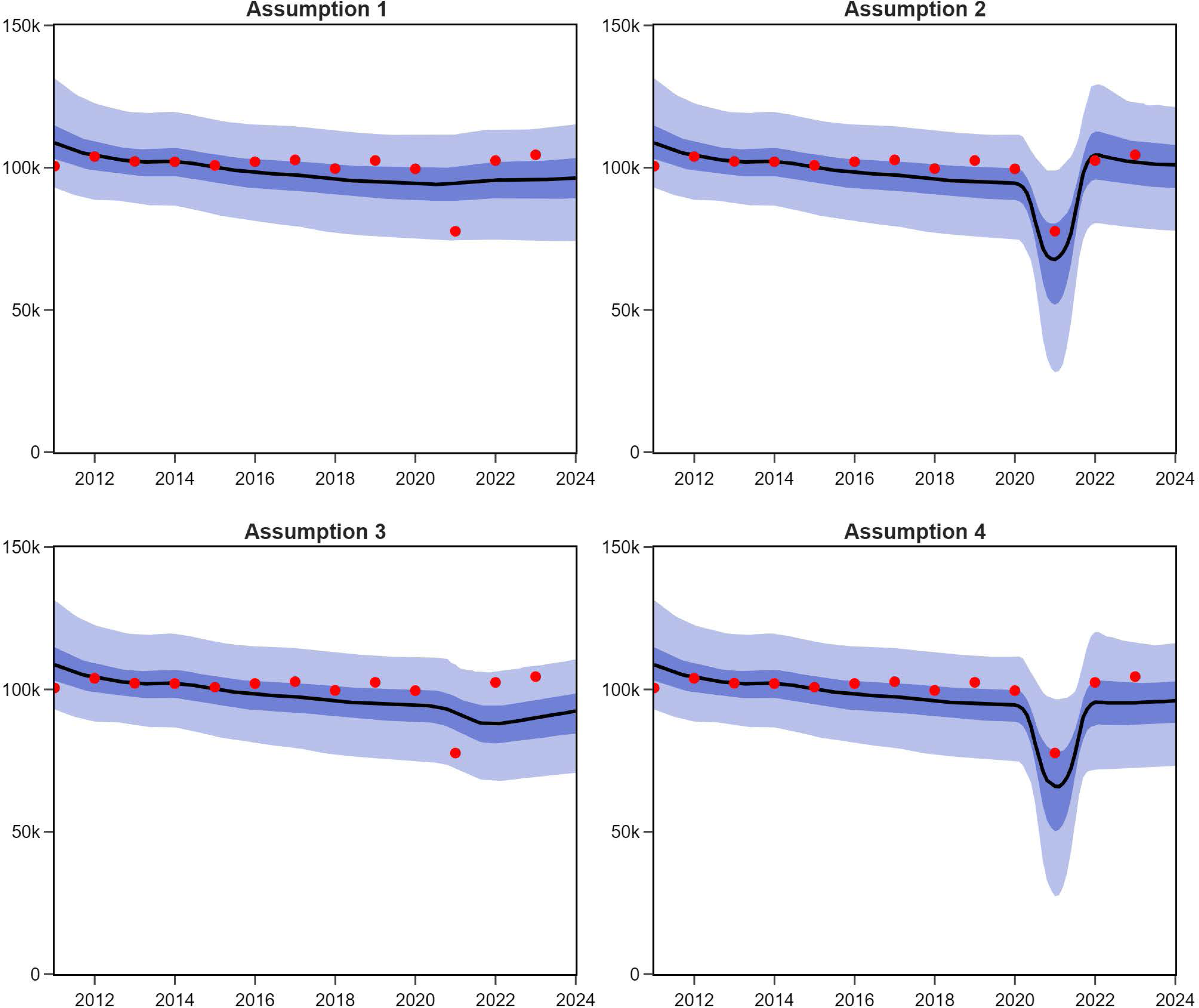
Model fits to the historical profile of case notifications under our four candidate assumptions regarding the effect of COVID-19 on *M.tb* dynamics. Solid lines represent median model estimates. Shaded areas show corresponding interquartile ranges (dark shade) and 95% credible intervals (light shade). Assumption 1: The COVID-19 pandemic had no impact on TB case detection or transmission. Assumption 2: Only case detection was reduced. Assumption 3: Only TB transmission was reduced. Assumption 4: Both case detection and TB transmission were reduced, incorporating effects from both social distancing and healthcare disruption. Alt text: Figure 2 shows the model’s fit to historical case notifications under four different assumptions about COVID-19’s impact on tuberculosis dynamics. The model’s median estimates are shown by solid lines, while the 95% credible intervals and interquartile ranges are shown by shaded areas (dark and light, respectively). Assumption 1, neither TB case detection nor transmission was impacted by the COVID-19 pandemic; Assumption 2, only case detection was decreased; Assumption 3, only TB transmission was decreased; and Assumption 4, decreases in both case detection and TB transmission.

Based on this analysis, we selected Assumption 2 as our primary model for the following in-depth examination of *M.tb* dynamics in Vietnam. Figure 3 illustrates the comparison between calibration targets and other epidemiological estimates not included in our calibration algorithm that we considered for validation. Figure S5 presents model outputs derived from 50 randomly accepted model runs.

**Figure 3.**
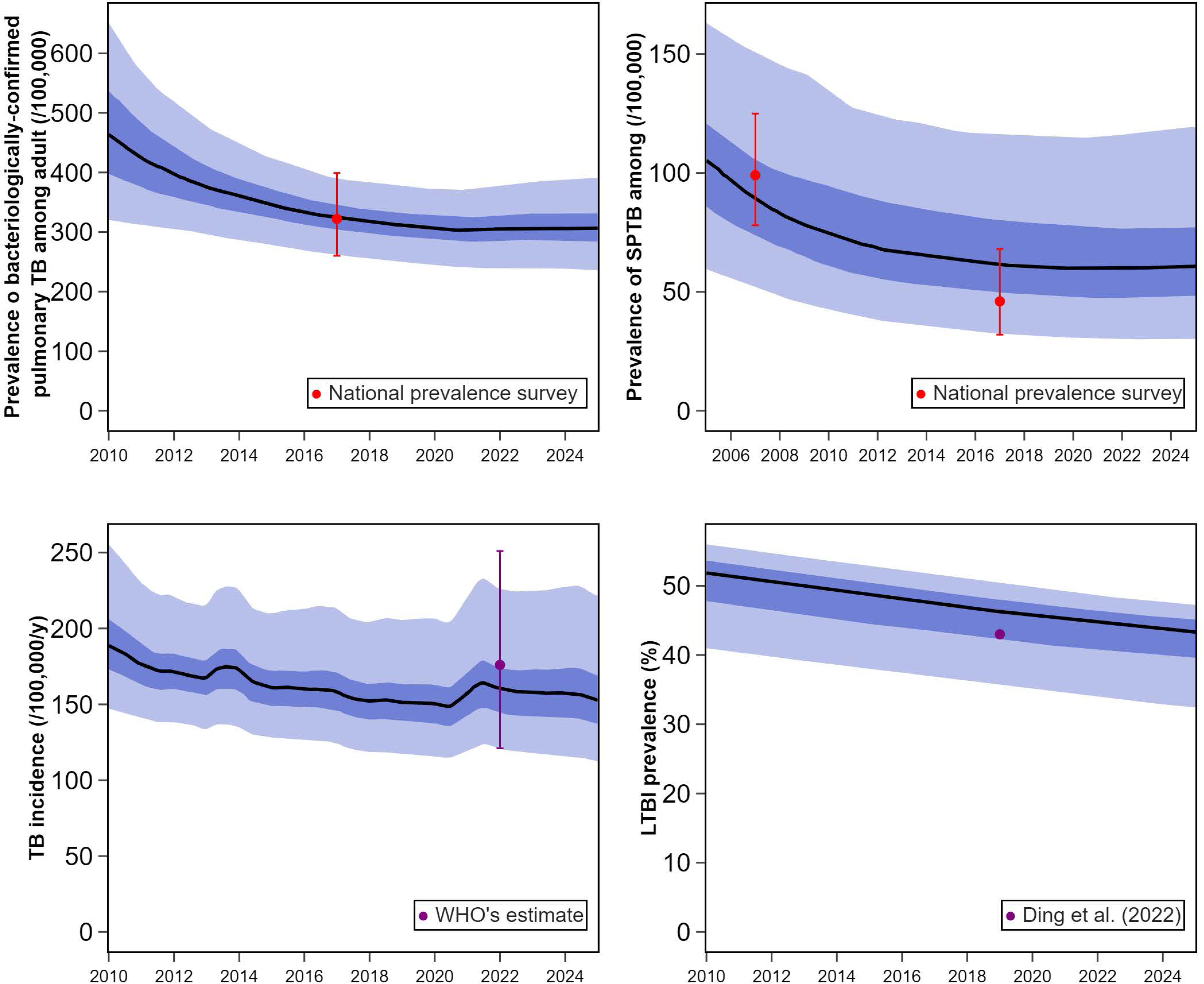
Comparison between outputs and empiric observation targets other than annual case notifications for Vietnam, under the best-fitting candidate model configuration (Assumption 2). Model predictions are represented as median (solid line), interquartile credible interval (dark blue shade) and 95% central credible interval (light blue shade). Red points and vertical line (where available) show the point estimates and confidence interval for each calibration target, while purple points indicate estimates from literature that are presented for comparison but were not used as calibration targets. Legends indicate the data source for each target. Alt text: Figure 3 illustrates a comparison between model outputs and empirical observation targets for Vietnam, excluding annual case notifications, under the best-fitting candidate model configuration (Assumption 2). The median is represented by a solid line, the interquartile credible interval by a dark blue line, and the 95% credible interval (CrI) by a light blue line. Red points with vertical lines that, when available, display the confidence intervals are used to indicate calibration targets. Estimates from the literature that are utilised for comparison but not as calibration objectives are shown by purple points. The legend provides specifics about each target’s data source.

Table S5 (Supplementary Material) details the posterior estimates and calibration metrics for the calibrated parameters of our preferred candidate model. The algorithm demonstrated good chain convergence, as evidenced by each parameter’s Gelman-Rubin statistic (*R*) being below 1.05. Figure S6 (Supplementary Material) shows the trace plots for each parameter, illustrating their convergence and stability throughout calibration. Figure S7 (Supplementary Material) overlays posterior distributions on the corresponding priors of the calibrated parameters. The posterior estimate for the reduction of COVID-19 on TB detection was 0.33 with 94% high density interval ranging from 0.01 to 0.68. Figure S8 (Supplementary Material) shows how the posterior distributions for the natural history parameters translate into the more epidemiologically intuitive quantities of disease duration and case fatality rate for SPTB and SNTB.

Our calibration algorithm simulated a peak in adult pulmonary TB prevalence that began to decline from around 1986, coinciding with the establishment of the NTP. By 2017, the model predicted an adult pulmonary TB prevalence of 326 per 100,000 populations (95%CrI: 264–387), closely matching our prevalence survey target, which estimated bacteriologically-confirmed TB among adults at 322 (confidence interval – CI: 260–399) per 100,000 in 2017 [11].

We estimated TB incidence in 2022 at 156,000, equivalent to 160 per 100,000 population (95%CrI: 122-218), which closely matches the WHO’s 2022 estimate of 176 per 100,000 (95%CI: 121–251, not used as a calibration target) [10]. We estimated that approximately 18,000 (18 per 100,000) people have died from TB each year since 2010. Our modelled prevalence of adult SPTB aligned closely with the results of the 2007 prevalence survey (90 per 100,000, 95%CrI: 53-153, compared to 99 per 100,000, 95%CI: 78-125) [17], but was higher than the 2017 survey results (63 per 100,000, 95%CrI: 32-119, compared to 46 per 100,000, 95%CI: 32-68) [11]. Figure S9 (Supplementary Material) illustrates the proportion of people with active TB originating from the early latent class, highlighting that 50% or more of these cases were due to early progression and therefore recent transmission throughout most of the recent simulation period. A minor increase in the proportional contribution of recent transmission was observed in 2021 during the COVID-19 pandemic. The modelled prevalence of latent TB infection (LTBI) in 2019 was 45% of the total population (95%CrI: 39%-51%). This aligns closely with the estimate from Ding et al. [18], which places the LTBI prevalence at 43.75% of the population. Figure S10 (Supplementary Material) presents the estimated new number of children developing TB per year. The model estimates that in 2023, approximately 26,800 children (95%CrI: 15,500–42,000) developed TB, accounting for 16% of total incidence – somewhat higher than the global level (12%) estimated by the WHO [19] and the figure of at least 7,800 previously mentioned by WHO for Vietnam [20]. In 2021, under Assumption 2, TB incidence rose by 3.1% in children compared to 1.8% in adults. Figure S11 (Supplementary Material) shows the proportions of the population by compartment based on results from the maximum likelihood run.

Figure S12 (Supplementary Material) shows the estimated increasing profile of the rate of commencing treatment over time, The rise began around 1986, coinciding with the establishment of the NTP and accelerated sharply in the late 1990s following DOTS implementation [20]. The median case detection proportion stabilized at around 0.6 from 2010 onwards, which can be interpreted as meaning that about 60% of new people with active TB were successfully detected and commenced treatment. Specifically, we estimated a 33% reduction in the rate of commencing treatment, during COVID-19, resulting in a 0.43-year (equivalent to 23 weeks) increase in the time to active TB onset to treatment.

### 2. Future projections

Figure 4 illustrates how the COVID-19-related health system effects may contribute to impeding future TB control efforts under Assumption 2, in comparison with a counterfactual scenario that COVID-19 had no effect on TB notifications. In 2021, we estimated 2,000 additional TB episodes (95%CrI: 200–5,100) and 1100 additional TB deaths associated with the effects of COVID-19 (95%CrI: 100–2,700). By 2035, the cumulative annual burden could rise by 22,000 TB episodes (95%CrI: 1,900–63,000) and 5,900 more deaths (95%CrI: 600–16,600). Figure S13 (Supplementary Material) shows the projected impact of the COVID-19 on cumulative TB episodes and TB-related deaths from 2021 to 2035 - under Assumption 2 and 4, compared to a counterfactual scenario with no COVID-19 effect on notifications and *M.tb* transmission. Figure S14 (Supplementary Material) presents estimates of the cumulative number of new TB episodes plotted against key model parameters, illustrating how variations in these parameters correlated with projected outcomes.

**Figure 4.**
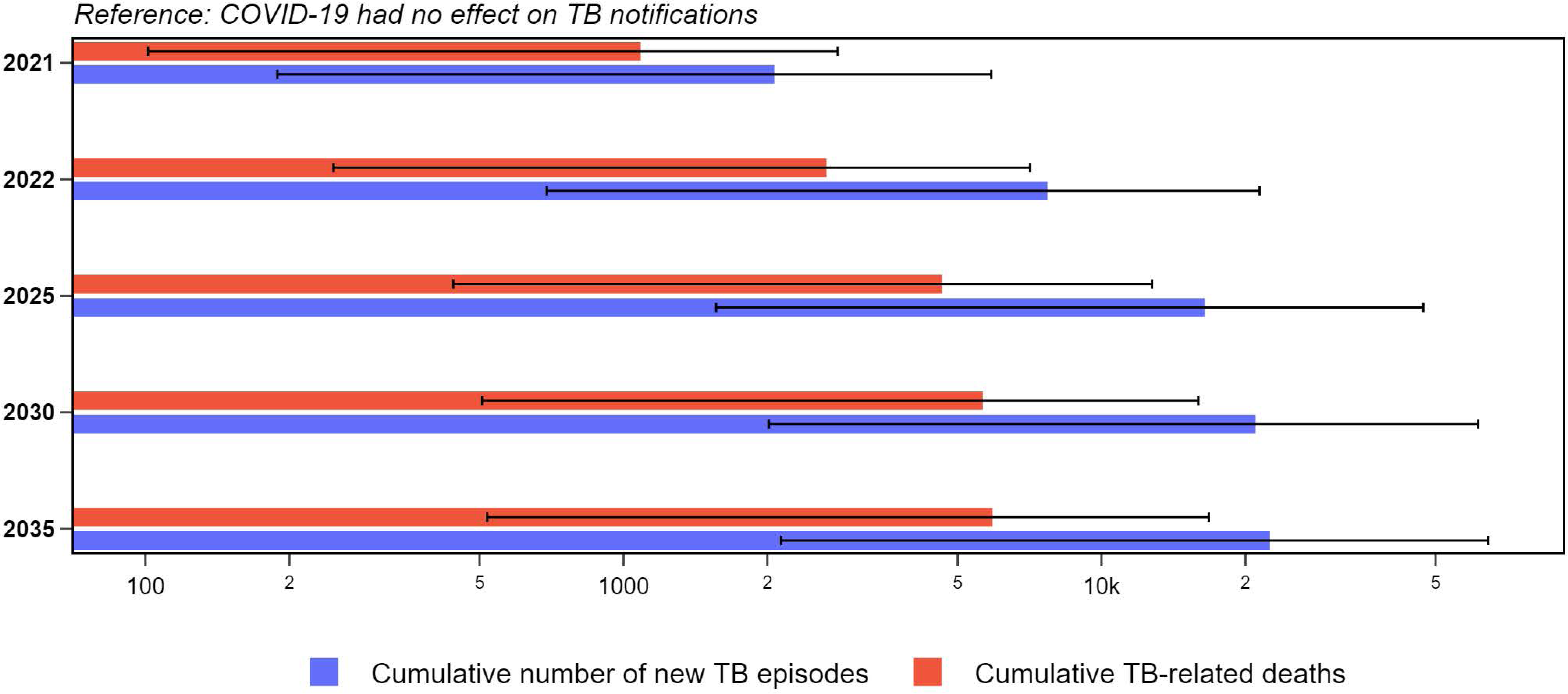
Impact of the COVID-19 pandemic on the cumulative number of TB episodes and TB-related deaths from 2021 to 2035 compared to a counterfactual scenario that COVID-19 had no effect on TB notifications. *Projections made under Assumption 2; values are presented on a logarithmic scale. Cumulative diseased: cumulative new TB episodes, Cumulative deaths: cumulative number of TB-related deaths. Lines indicate the 95% credible intervals of the estimates*. **Alt text:** Figure 4 displays the impact of the COVID-19 pandemic on the cumulative number of tuberculosis (TB) episodes and TB-related deaths from 2021 to 2035, compared to a counterfactual scenario where COVID-19 had no effect on TB notifications. Assumption 2 is used to make the projections, and a logarithmic scale is used to display the data. The graph labels “Cumulative deaths” and “Cumulative diseased” denote the total number of TB-related deaths and cumulative new TB cases, respectively. The graph’s lines show the estimations’ 95% credible intervals.

In 2024, we estimated the average delay to diagnosis and treatment for a person with TB at 1.80 years, such that our scenarios are equivalent to reducing the delay to 0.9 years (47 weeks) with a 2-fold increase, and to 0.36 years (19 weeks) with 5-fold in rate of commencing treatment. Our projections of the results of these scenarios are presented in Figure 5. Compared to a *‘status quo’* projection, enhanced case detection by increasing the rate of commencing care 5-fold could prevent approximately 434,000 (95%CrI: 250,000–692,000) new TB episodes, representing a 17.8% reduction (95%CrI: 13.1%-21.9%). Additionally, this scenario may avert 110,000 (95%CrI: 69,000–170,000) TB-related deaths, equivalent to 34.2% (95%CrI: 31.5%-37.0%) of all TB-related deaths over the coming decade. Notably, enhancing TB case detection may have an even greater impact on reducing TB-related mortality.

**Figure 5.**
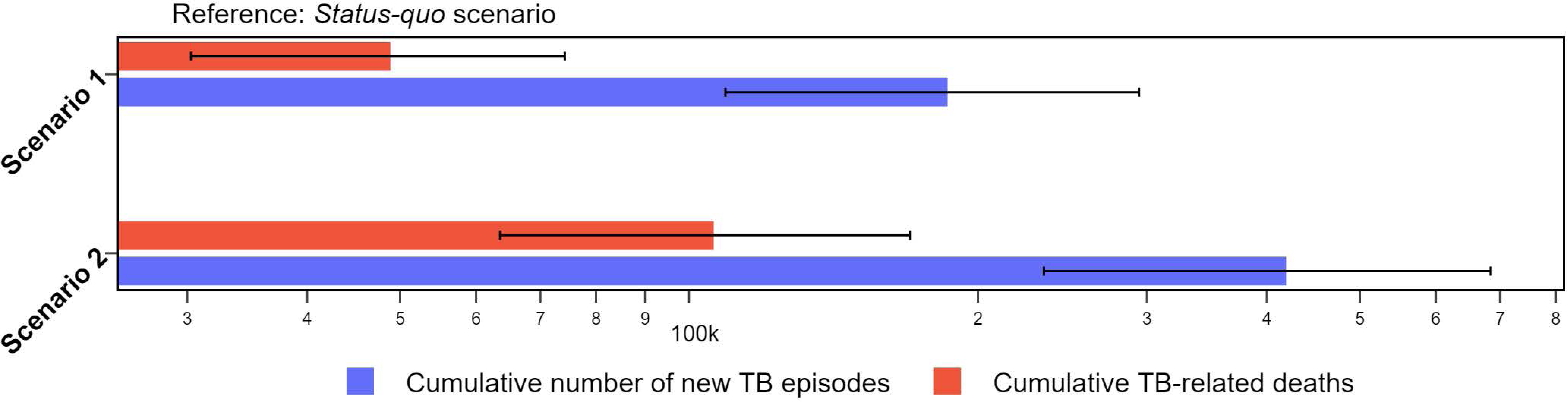
Number of cumulative new TB episodes and TB-related deaths averted under two scenarios of enhancing case detection comparing to *‘status quo’ scenario*. Projections made under Assumption 2, values are presented on a logarithmic scale. The ‘status quo’ scenario assumes case detection remains unchanged. Scenario 1 models a 2-fold increase and Scenario 2 models a 5-fold increase in the rate at which persons with active TB commence care. Lines indicate the 95% credible intervals of the estimates. Alt text: The number of cumulative new TB episodes and TB-related deaths prevented under two scenarios that improve case detection in comparison to a “status quo scenario” is displayed in Figure 5. The results are displayed on a logarithmic scale, and the projections are made under Assumption 2. In the ‘status quo’ scenario, case detection is assumed to stay constant. The rate at which people with active TB start receiving care is predicted to increase by two times in Scenario 1 and five times in Scenario 2. The graph’s lines show the 95% credible intervals of the estimates.

## DISCUSSION

Our results show that the decline in TB detection during COVID-19 was primarily responsible for the sharp drop in TB notifications in Vietnam in 2021. This disruption is projected to result in long-term increases in TB incidence and mortality, extending long beyond the period of disruption. However, enhancing case detection could substantially mitigate these long-term impacts and accelerate progress by reducing both incidence and mortality in the years following the pandemic. The novelty of our study lies in the integration of dynamic transmission modeling with a flexible, modular framework built using open-source softwares. Our approach allows for the simulation of pandemic-related disruptions and future intervention scenarios, while supporting reproducibility and adaptability [16].

The close alignment between modelled outputs and calibration targets demonstrates our model’s capacity to integrate multiple key observations reflecting TB burden with empirically derived natural history parameters. We found that COVID-19 was associated with significant reductions in TB detection, which poses a serious threat to long-term TB control efforts and aligns with findings from other studies [21, 22]. However, Vietnam has adapted its policies in light of the lessons of the COVID-19 pandemic and is working to enhance TB detection, focusing on decentralized services In response to COVID-19, Vietnam has strengthened TB detection through decentralized services and the ‘Double X’ strategy - combining chest radiography with GeneXpert - to improve early diagnosis among vulnerable populations [23–25]. Continuing and expanding these programs may further strengthen TB prevention and care, and mitigate long-term disruptions to TB control efforts.

The assumption incorporating reduced case detection aligned most closely with observed trends, suggesting that COVID-19-related disruptions likely delayed TB diagnosis and treatment, leading to fewer reported cases. While we cannot exclude that reduced contact may have contributed slightly, the marked dip in case detection likely had a greater impact. The resulting backlog of undiagnosed cases may continue to drive transmission.

The modelled delay from active TB onset to treatment can be interpreted as being exponentially distributed with an average time to treatment of approximately 1.80 years, implying that many people with TB remain undetected. This delay may appear higher than prior studies which reported median diagnostic delays of 28 days in low- and lower-middle-income countries and 10 days in upper-middle-income countries [26]. In high-endemic areas, delays often exceeded 120 days, influenced by gender, rural residence, and health system inefficiencies [26, 27]. Variation in the definition of first healthcare-seeking behavior further complicates comparisons, with definitions ranging from visits to qualified providers to contact with the NTP [26]. The long delay can be partly explained by the high prevalence and prolonged disease duration of SNTB. Despite advances in molecular diagnostics, SNTB remains difficult to detect due to mild or absent symptoms [28].

The wide uncertainties in our projections reflect the fact that disruptions to TB case detection are not directly measurable. Surveillance systems report only diagnosed cases, offering no visibility into those who developed TB but were never detected or treated. As a result, the extent of missed or delayed diagnoses during the pandemic must be inferred indirectly, contributing to the uncertainty in estimation and future projection. Even so, these uncertainty ranges can guide policy. Policy-makers should act on the median estimate but remain prepared for worse-case outcomes. Interventions that are beneficial across all scenarios - such as expanded case-finding and improved diagnostic access - should be prioritized. These findings highlight the urgent need to restore and strengthen TB detection systems to mitigate long-term consequences, which requires sustained investment and commitment. Future disruptions - whether pandemics or other crises - could further jeopardize TB prevention and care.

Our study has limitations stemming from data availability and from uncertainties in TB epidemiology and natural history. First, the model candidates were calibrated to the best available observed data, including TB notifications data from the NTP and results from the national prevalence survey. However, TB notifications may still be affected by incomplete capture of treated cases in surveillance data or false-positive diagnoses. This limitation is particularly pronounced in children, for whom diagnosis is more challenging and bacteriological confirmation rates are low, such that current approaches may not fully capture the true disease burden in this population. Additionally, changes in case definitions over time, driven by the shift from smear microscopy to more sensitive microbiological diagnostics, may influence trends in notifications and complicate interpretation of historical data.

Second, while recent research provides estimates for progression rates from LTBI to active TB, LTBI reactivation remains difficult to observe [29, 30]. We relied on several estimates from low-burden settings or from the pre-chemotherapy era that may not be fully applicable to the context of Vietnam. The difficulty in conducting longitudinal studies in high-burden areas further limits the precision of our estimates, potentially affecting the generalizability of our findings [31]. Despite this, these estimates aligned well with observed data, and our infection natural history parameters were empirically grounded, providing greater confidence than unsubstantiated assumptions such as a 5-10% lifetime risk [30]. Uncertainties in input parameters likely contributed to the wide range of future projections. We conceptually aligned our natural history categories with the historical “open” vs. “closed” TB distinction - roughly corresponding to more infectious, symptomatic (SPTB) and less infectious or extrapulmonary forms. While this is imprecise and may not correspond directly to smear-based diagnostic status, this mapping broadly reflects current clinical understanding and incorporates some heterogeneity in disease forms. The difficulty in conducting longitudinal studies in high-burden areas further complicates the precision of these estimates [31]. The major community-wide active case finding trial in Ca Mau province [32] provides opportunities for better understanding TB epidemiology in Vietnam and our modelling analysis can be considered in this context. The significant prevalence reduction observed during the trial period may suggest early progression may contribute even more to people with TB than late reactivation and our model estimated. This potential underestimation highlights an important area for future research to further refine *M.tb* transmission dynamics.

Third, our model implemented a synthetic social contact matrix from 2007, and an updated matrix would more accurately reflect current transmission dynamics, considering changes in social behavior, contact patterns, and demographics. The matrix reflects average population contact patterns but may overlook setting-specific transmission, such as within households or close-proximity transmission during COVID-19 quarantine.

Fourth, although we assumed that the average delay to diagnosis and treatment initiation would not change over the next ten years in our *“status-quo”* scenario, future changes - such as expansion of NTP capacity, which could shorten this delay, or reductions in funding, which could compromise treatment coverage - may alter this trajectory. Moreover, while important, addressing drug-resistant TB was beyond the scope of this study.

Our study’s strengths include our calibration approach, which harnessed open-source software and successfully captured key indicators of the epidemic while also reflecting uncertainties around key input parameters. The model was informed by multiple relevant data streams, including results from the recent TB national survey. This allowed us to replicate the local TB burden and ensure that historical trends in case detection were reflected through TB notifications. Although the model was not calibrated to WHO’s estimates of TB incidence and mortality - given they are derived from modelling rather than surveillance - our results closely align with those estimates. Our pipeline can simulate a broad range of directly transmitted pathogens, enables accurate, transparent with built-in support for calibration and uncertainty analysis [16].

## CONCLUSIONS

Our study underscores the significant impact of COVID-19-related disruptions on TB case detection in Vietnam, leading to a backlog of undiagnosed people with TB that could sustain transmission for years. However, these impacts may be mitigated by enhancing case detection activities, which can improve early detection and reduce transmission risks. Any major disruption now or into the future - whether due to another pandemic, other public health crises - could further exacerbate TB prevention and care challenges. Addressing these risks requires sustained investment in diagnostic capacity, healthcare system resilience, and proactive policies to mitigate the long-term consequences of such disruptions on TB prevention and care. Although we initially hypothesised that COVID-19 disruptions might provide insights into the underlying dynamics of the TB epidemic, they ultimately revealed the considerable uncertainty in assessing TB burden, which remains largely inferred from a single annual notification count. to mitigate the long-term consequences of such disruptions on TB prevention and care.

## DECLARATIONS

### Ethics approval and consent to participate

Not applicable, as this study involves modeling and does not include human participants or identifiable data.

### Availability of data and materials

The data supporting this article and the code used for the analyses are accessible in a GitHub repository at: https://github.com/vlbui/tbdynamics.

### Author Contributions

**Viet Long Bui:** Conceptualization, Methodology, Software, Data Curation, Formal analysis, Writing- Original draft preparation; Software; Writing- Reviewing and Editing; **Romain Ragonnet:** Conceptualization, Methodology, Software, Validation, Formal analysis, Writing- Reviewing and Editing; **Angus E. Hughes:** Methodology, Writing- Reviewing and Editing; **David S. Shipman:** Software; **Emma S. McBryde:** Writing- Reviewing and Editing; **Binh Hoa Nguyen:** Writing- Reviewing and Editing; **Hoang Nam Do:** Writing- Reviewing and Editing; **Thai Son Ha:** Writing- Reviewing and Editing; **Greg J. Fox:** Conceptualization, Data Curation, Methodology, Writing- Reviewing and Editing; **James M. Trauer:** Conceptualization, Methodology, Software, Validation, Formal analysis, Writing- Reviewing and Editing, Supervision.

## Acknowledgements

Viet Long Bui is a recipient of a postgraduate scholarship from Monash University. James M. Trauer is the recipient of a Discovery Early Career Fellowship from the Australian Research Council (DE230100730). The funders had no role in study design, data collection and analysis, decision to publish, or preparation of the manuscript.

