## Supplementary Material for "Impact of Case Detection and COVID-19-Related Disruptions on Tuberculosis in Vietnam: A Modelling Analysis"

**Table of** **Contents**

### I. Model structure

#### 1. Compartments

The model's compartments were:

- Susceptible (S): Individuals who have never previously been infected with *M.tb* and are at risk of initial infection.
- Early latent infection (E): People in this category have recently contracted *M.tb*, but have not yet developed features of active TB disease. They face a heightened risk of developing active TB through this early period following their initial infection.
- Late latent infection (L): This category represents persons who have had latent TB infection for a longer period than for those in ‘E’. Their likelihood of progressing to active TB compared to those in the early latent phase has declined through immune containment of the pathogen.
- Active TB (I): Individuals in this compartment have active TB disease and can spread the disease to others in the categories that can be infected (S, L and R).
- Treatment (T): Individuals in this compartment are currently undergoing treatment with anti-tuberculous chemotherapy.
- Recovered (R): This compartment represents those who have previously recovered from active TB disease, either through natural resolution or through completing treatment. These individuals may be reinfected and transition back into the compartment (E).

##### 1.1 Stratification by age

We divided our model into six sequential age groups to represent the population aged 0-4, 5-14, 15-34, 35-49, 50-69, and 70+ years. As well as being used to capture the demographic processes of birth, ageing and death, this population partition allowed for the consideration of differential interaction behaviours among each pair of age groups via an age-specific interaction matrix. To simulate this heterogeneous mixing across age groups, we used a contact matrix (Figure S1) adapted to these age categories, drawing on data from a contact survey conducted in semi-rural areas of Viet Nam [1].

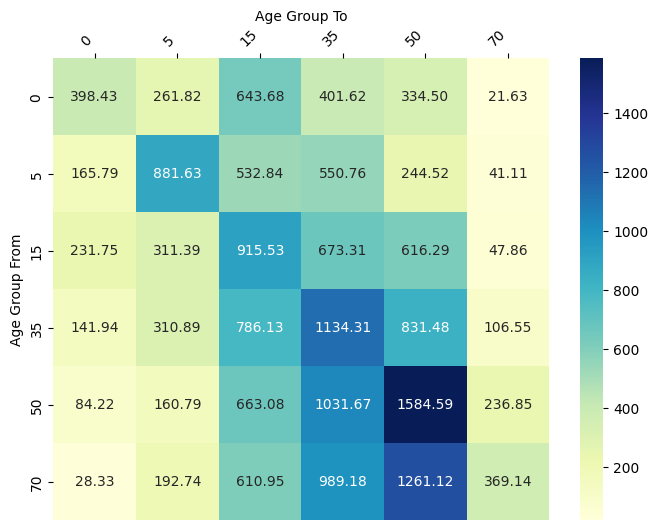

**Figure S1. Yearly contact matrix across age groups.**

The matrix illustrates the average number of contacts per year between different age groups. The rows represent the age groups of individuals reporting contact, while the columns represent the age groups of individuals they contacted. The values in each cell reflect the yearly total number of contacts between individuals in the corresponding age groups. The code used to generate the contact matrix is available at <https://github.com/monash-emu/AuTuMN/blob/master/autumn/tools/inputs/social_mixing/build_synthetic_matrices.py>).

##### 1.2. Stratification by pulmonary/smear status

Our model captures three distinct clinical manifestations of TB, categorized according to the patient’s organ involvement and smear status. Specifically, we included categories for smear-positive TB (SPTB), smear-negative pulmonary TB (SNTB), and extrapulmonary TB (EPTB).This stratification was incorporated to allow the active disease compartment (I) to exhibit varying levels of disease severity, infectiousness, and rates of detection in the flow from (I) to treatment (T).

**Table S1. Summary of stratification effects**

| **Stratifications** | **Strata** | **Rationale for inclusion and modelled effects** |
| --- | --- | --- |
| Age group | - 0-4 years-old - 5-14 years-old - 15-34 years-old - 35-49 years-old - 50-69 years-old - 70 years-old and over | - Heterogeneous mixing by age. - Variable risk of progression from latent to active TB by age group. - Variable background (non TB-related) mortality rates by age group. - Variable age-specific infectiousness. - Variable BCG vaccine effect and coverage by age group. |
| Clinical form of TB | - SPTB - SNTB - EPTB | - Variable case detection rates by organ involvement status. - Higher infectiousness for SPTB than for SNTB. - EPTB considered non-infectious. |

##### 1.3. Demographics

In the model, births are introduced through time-varying crude birth rates, which are calculated from the total population size to determine the rate at which newborns enter the model over time. Mortality unrelated to TB is accounted for using time-varying, age-specific mortality rates that apply uniformly across all compartments. These birth and non-TB-related mortality rates are estimated from data from the United Nations Population Division [2] (UN Population). We also applied additional TB-specific mortality rates to the (I) and (T) compartments, ensuring that deaths directly attributable to TB disease were accurately reflected.

#### 2. *M.tb* transmission

In the context of *M.tb* transmission, our model distinguished between varying levels of susceptibility among individuals based on their prior exposure to *M.tb*. Those who are latently infected or have previously recovered from active TB exhibit a different level of susceptibility to infection compared to those who have never been infected.

We incorporated the protective effect of BCG vaccination by reducing susceptibility to infection for individuals younger than 30 years, aligning with the age groups likely to have received routine BCG immunisation in Viet Nam since the 1980s. While BCG has demonstrated clear protective effects in children and early adolescents, the extent to which this protection persists into adulthood is less well established [3]. Prior studies, including a retrospective cohort study in Norway, have shown that BCG effectiveness may decline with time since vaccination, whereas earlier trials by the UK Medical Research Council (MRC) reported sustained protection into early adulthood following childhood vaccination. We assumed a 70% reduction in susceptibility for BCG-vaccinated children under 15 years old [4]. Instead of using an explicit stratification of compartments, we used a linear model to represent the gradual decrease in BCG vaccine immunity from age of 15 to 30 (Figure S2). We modelled BCG-induced immunity as a step function across age bands, incorporating average waning effects and adjusting susceptibility based on WHO-estimated fluctuations in BCG coverage in Viet Nam, as estimated by the World Health Organization (WHO). Importantly, the latency parameters used in our model - derived from cohorts with low BCG coverage in high-income settings - would otherwise imply very high (and likely unrealistic) early progression rates in children (e.g.30 - 40% progression to active TB within six months of exposure [5]). Without incorporating BCG’s protective effect, we would therefore have overestimated childhood TB incidence or needed to parameterise our model using an alternative approach that was not empirically derived. Our implementation is therefore essential to reflect a more realistic age-specific disease burden, consistent with previous modelling approaches. Our approach also captures greater historical realism over the course of BCG coverage expansion during the 20^th^ Century and incorporates uncertainty in the vaccination-related parameters without the need to increase the number of model compartments. An identical approach was used previously by Ragonnet et al. [6].

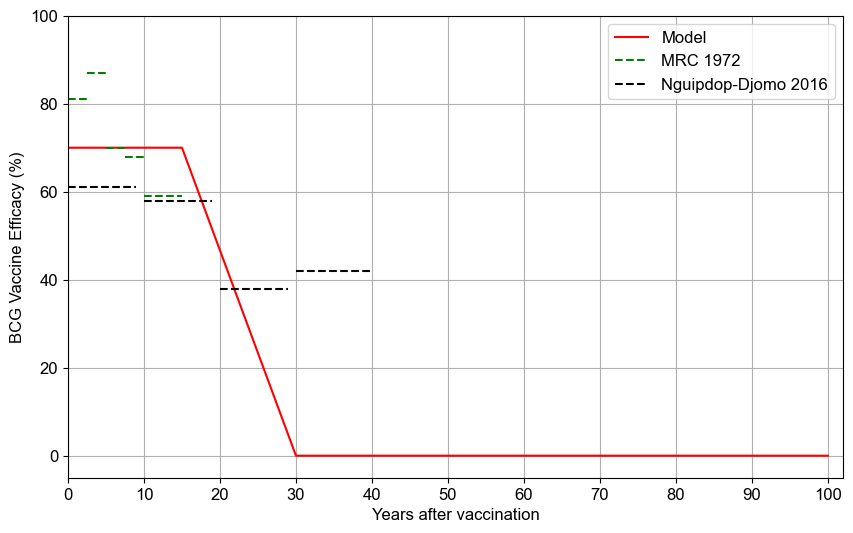

**Figure S2. Assumed waning profile of BCG efficacy.** *Green and black dashed lines represent estimates of Medical Research Council (MRC)* [7] *and Nguipdop -Djomo et al.* [4]*, while the red line shows the modelled vaccine effect.*

We assumed that SNTB was 25% as infectious as SPTB, while EPTBTB was modelled as a non-infectious disease state [8,9].

Infectiousness was assumed to be higher for adults (age ≥15 years), and we used a logistic function $age=\frac{1}{1+e^{-\left( age-15 \right)}}$ to model a progressive increase with age [6,10]. Finally, individuals who are on treatment (T compartments) were assumed to be partially infectious, as infectiousness declines rapidly after treatment initiation. Infectiousness was multiplied by 0.08 for the treatment compartment compared to the untreated disease compartment to reflect that individuals may remain infectious for around two weeks out of the 26 weeks of a standard regimen [11].

##### 2.1. Latency and disease progression natural history parameters

We used previously reported empirical estimates to inform the dynamics of the progression from infection to active TB disease [12]. These parameters, which guide the model's representation of the transition from infection to disease, vary with the age of the modelled individuals. Moreover, because of the importance of these parameters, to accommodate the uncertainties inherent in these progression rates, we incorporated a multiplier as undertaken in [12].

We used estimates from the pre-chemotherapy era reported by Ragonnet et al. to model rates of TB mortality and self-recovery in the absence of treatment [13]. We employed different rates of untreated TB mortality and self-recovery for SPTB compared to SNTB. The TB mortality and self-recovery rates associated with EPTB were assumed to be the same as those of SNTB.

##### 2.2. Passive detection of active TB disease

We represented passive detection as the rate at which individuals with active TB (I) transition to the treatment (T), or simply the rate of commencing treatment from active TB. We considered this rate as the product of: the reciprocal of the average time from developing active TB to first presentation and treatment initiation under the National Tuberculosis Program (NTP); and the diagnostic algorithm's sensitivity, accounting for the relative difficulty diagnosing SNTB and EPTB compared to SPTB.

We modelled the rate of treatment commencement as increasing over time, consistent with our historical understanding of the epidemic and the fact that TB treatment has only been available for a number of decades in Viet Nam. In particular, improvements in treatment followed the establishment of the NTP in 1986 and the implementation of the Directly Observed Treatment, Short-Course (DOTS) strategy from 1992 [14]. These initiatives significantly improved case detection by enhancing diagnostic methods, leading to a steady rise in case notifications. To capture this progression, we used a smooth transition function that gradually increased over this period of improving access to TB diagnostics and treatments. The function is governed by input parameters that determine the timing of the maximum gradient, which represents the point of the most rapid change in detection rates, the peak rate of increase, and the peak average delay from the onset of active TB to diagnosis. These parameters were calibrated to align with observed data, ensuring that the model accurately reflects real-world trends in TB detection dynamics.

##### 2.3. Treatment outcomes

Once diagnosed and commenced treatment, an individual with active TB will be treated regardless of their pulmonary or smear status. In our model, treated individuals can experience three distinct treatment outcomes: treatment success, relapse, and death. We do not explicitly model sputum smear conversion. Treatment outcomes are calculated from the following parameters: $\Pi$ (proportion of non-successful outcomes in treatment that result in death), $\mu$ (natural death rate), $T$ (treatment duration), $S$ (treatment success proportion, often referred to as the ‘treatment success rate’). The proportion of non-TB-related deaths occurring during treatment is given by: $1-e^{-\mu T}$. The proportion of deaths resulting from TB during treatment is given by: $\left( 1-S \right)\times\Pi$. The proportion of deaths attributable to treatment is calculated by subtracting the proportion of natural deaths occurring during the treatment period from the total proportion of expected deaths, while ensuring the result is non-negative.

The proportion of treatment episodes resulting in relapse was determined by subtracting the adjusted death rate and the natural death proportion from 1 minus the treatment success proportion. Finally, the rates of transition for use in the model were obtained by multiplying each proportion by $T$, providing adjusted values for success, death, and relapse.

#### 3. Model equation

The force of infection $\lambda_{a}\left( t \right)$, representing the hazard of infection for susceptible individuals in age group $a$, is defined as:

$$\lambda_{a}\left( t \right)=\sum_{a'} C_{a,a'}\sum_{f} \epsilon_{f}\frac{I_{a',f}\left( t \right)+\eta T_{a'}\left( t \right)}{N_{a'}\left( t \right)}$$

where:

- $C_{a,a'}$: Contact rate between age groups $a$ and $a'$
- $\epsilon_{f}$: Relative infectiousness of TB form $f$
- $\eta$: Relative infectiousness during treatment
- $N_{a'}\left( t \right)$: Total population in age group $a'$ at time $t$

The age-structured model is defined by the following system of differential equations:

$$\begin{matrix} \frac{dS_{a}}{dt} & =B_{a}\left( t \right)+A_{a-1\to a}\left( S \right)-A_{a\to a+1}\left( S \right)-\lambda_{a}\left( t \right)S_{a}-\mu_{a}S_{a} \\ \frac{dE_{a}}{dt} & =\lambda_{a}\left( t \right)\left[ S_{a}+\kappa_{L}L_{a}+\kappa_{R}R_{a} \right]+A_{a-1\to a}\left( E \right)-A_{a\to a+1}\left( E \right)-\left( \sigma_{a}+\rho_{a}+\mu_{a} \right)E_{a} \\ \frac{dL_{a}}{dt} & =\sigma_{a}E_{a}+A_{a-1\to a}\left( L \right)-A_{a\to a+1}\left( L \right)-\left( \gamma_{a}+\mu_{a} \right)L_{a} \\ \frac{dI_{a,f}}{dt} & =\pi_{f}\left( \rho_{a}E_{a}+\gamma_{a}L_{a} \right)+A_{a-1\to a}\left( I_{f} \right)-A_{a\to a+1}\left( I_{f} \right)-\left( \theta_{a,f}+\mu_{TB,a,f}+\mu_{a}+\gamma_{f} \right)I_{a,f} \\ \frac{dT_{a}}{dt} & =\sum_{f} \theta_{a,f}I_{a,f}+A_{a-1\to a}\left( T \right)-A_{a\to a+1}\left( T \right)-\left( \psi+\delta+\mu_{a} \right)T_{a} \\ \frac{dR_{a}}{dt} & =\psi T_{a}+\sum_{f} \gamma_{f}I_{a,f}+A_{a-1\to a}\left( R \right)-A_{a\to a+1}\left( R \right)-\lambda_{a}\left( t \right)\kappa_{R}R_{a}-\mu_{a}R_{a} \end{matrix}$$

where:

Compartments (all stratified by age group $a$ and time $t$):

- $S_{a}\left( t \right)$: Susceptible individuals
- $E_{a}\left( t \right)$: Early latent TB infection
- $L_{a}\left( t \right)$: Late latent TB infection
- $I_{a,f}\left( t \right)$: Active TB (form $f$)
- $T_{a}\left( t \right)$: On TB treatment
- $R_{a}\left( t \right)$: Recovered (post-treatment or self-recovery)

Demographic and infection dynamics:

- $B_{a}\left( t \right)$: Births into age group $a$ (non-zero only for youngest group)
- $A_{a-1\to a}\left( \cdot\right)$, $A_{a\to a+1}\left( \cdot\right)$: Ageing flows
- $\mu_{a}$: Natural mortality rate for age group $a$
- $\lambda_{a}\left( t \right)$: Force of infection for age group $a$
- $N_{a'}\left( t \right)$: Total population in age group $a'$

Infection progression and reinfection:

- $\kappa_{L}$: Relative risk of reinfection while latently infected
- $\kappa_{R}$: Relative risk of reinfection after recovery
- $\sigma_{a}$: Stabilization rate from early to late latency (age-specific)
- $\rho_{a}$: Rate of rapid progression to active TB (age-specific)
- $\gamma_{a}$: Rate of late reactivation from latency (age-specific)

Clinical form and transmission:

- $\pi_{f}$: Proportion of new TB assigned to form $f$
- $\epsilon_{f}$: Relative infectiousness of form $f$
- $\eta$: Relative infectiousness during treatment

Treatment and outcomes:

- $\theta_{a,f}$: Treatment initiation rate (age and form-specific)
- $\mu_{TB,a,f}$: TB-specific mortality (age and form-specific)
- $\gamma_{f}$: Self-recovery rate for form $f$
- $\psi$: Treatment success rate
- $\delta$: TB-related mortality during treatment

### II. Parameterisation and calibration

#### 1. Parameterisation

Table S6 provides an overview of the model parameters. Several key parameters, identified as critical to model performance, will be varied during the calibration process.

#### 2. Model implementation and calibration

**2.1. Model Implementation**

We initiated the model in the year 1800 with TB absent, using a time step of 0.1 years to allow demographic dynamics to stabilise independently before TB was introduced in 1805 by adding one individual with active, transmissible disease through a short, time-limited pulse implemented as a triangular wave function..

Model parameters are detailed in Table S2, and calibration targets were set to reflect observed overall population size, annual reported TB notification and results from national TB surveys (prevalence of pulmonary TB among adults, prevalence of SPTB among adults) (Table S3).

**Table S2. Model parameters**

| **Parameter** | **Description** | **Value/Prior distribution** | **Evidence/Rationale** |
| --- | --- | --- | --- |
| Crude birth rate | The per capita rate at which new individuals are born into the population each year. | Time-variant | United Nations population [2] |
| Universal death rate | The per capita rate of mortality from causes unrelated to TB, applied across all compartments based on age group and calendar time. | Time-variant | United Nations population [2] |
| Contact rate | The per capita rate of potentially infectious contacts per year, governing the force of infection in the population. | Uniform distribution [15], range: 0.001 – 0.05 | Assumed. Minimally informative prior with bounds spanning the plausible range of values. |
| Relative infectiousness of SPTB/SNTB/EPTB | The infectiousness of individuals with SPTB, SNTB, and EPTB, expressed as a proportion relative to SPTB TB (baseline = 1.0). SNTB is assumed to be less infectious, and EPTB is assumed non-infectious. | 1 / 0.25 / 0 | The relative transmission rate of SPTB compared with SNTB patients has been estimated to be approximately one-quarter [8,9] |
| Relative infectiousness of adults (≥15 years) to children (<15 years) | A scaling factor representing how infectiousness increases with age, with adults assumed to be more infectious than children. Modelled using a logistic function that is then discretised to the modelled age groups. | Progressive increase through childhood using a logistic function, capturing its nonlinear development with an inflection point at age of 15. | Infectiousness was assumed to be higher for adults [16] |
| Infectiousness during treatment (relative to untreated TB) | Multiplier applied to infectiousness while undertaking treatment, reflecting that treated individuals remain infectious for only a short period after treatment begins. | 0.08 | Based on the assumption that patients are infectious for the first 2 weeks of a 6-month regimen [11]. |
| Rate of stabilization from early to late latency, per year | The annual per capita rate at which individuals with early latent TB transition to late latency, representing a more stable infection state with a lower risk of progression to active disease. | Age 0-4: 4.4 per year Age 5-14: 4.4 per year Age 15+: 2 per year | Maximum likelihood estimates of age-specific daily rates of stabilization from Table 1 of [12] converted to yearly rates. |
| Rate of rapid progression to active TB, per year | The annual per capita rate at which individuals with a new TB infection rapidly progress to active TB disease, bypassing the latent phase. | Age 0-4: 2.4 per year Age 5-14: 1 per year Age 15+: 0.1 per year | Maximum likelihood estimates of age-specific daily rates of stabilization from Table 1 of [12] converted to yearly rate. |
| Rate of late reactivation | The annual per capita rate at which individuals with latent TB infection progress to active TB disease after a prolonged period of dormancy. | 7e-9 per year Age 5-14: 2.3e-3 per year Age 15+: 1.2e-3 per year | Maximum likelihood estimates of age-specific daily rates of stabilization from Table 1 of [12] converted to yearly rate. |
| Uncertainty multiplier for the rates of late reactivation | A multiplier applied to the base rate of late reactivation, allowing the model to account for uncertainty in the long-term risk of reactivation. | 0.5 - 2 | Assumed and consistent with the approach of [12] |
| Proportion of pulmonary TB among incidence | The proportion of all incident TB cases that are pulmonary (as opposed to extrapulmonary). | Uniform distribution, range: 0.5 – 0.95 | Assumed to replicate the proportion observed in the NTP data. |
| Proportion of SPTB among pulmonary incidence | The proportion of pulmonary TB cases that are SPTB, which are the most infectious form of TB. | Uniform distribution, range: 0.6 – 0.95 | Assumed based on typical ranges observed in national TB survey; SNTB are a substantial proportion of pulmonary TB. |
| SPTB mortality, per year* | The annual per capita rate at which individuals with SPTB die from TB in the absence of treatment. | Truncated normal distribution [17], standard deviation (SD): 0.028, range: 0.335 - 0.449 | - Maximum likelihood estimates of SPTB mortality from Figure 3 of [13].  - To prevent sharp cut-offs at the boundaries while still capturing the plausible range of values. |
| SPTB self-recovery rate, per year* | The annual per capita rate at which individuals with SPTB recover from TB without receiving treatment. | Truncated normal distribution, range: 0.177 - 0.288, SD: 0.029. | As preceding parameter |
| SNTB mortality, per year* | The annual per capita rate at which individuals with SNTB die from TB in the absence of treatment. | Truncated normal distribution, range: 0.017 - 0.035, SD: 0.004. | - Maximum likelihood estimates of SNTB mortality from Figure 4 of [13].  - To prevent sharp cut-offs at the boundaries while still capturing the plausible range of values. |
| SNTB self-recovery rate, per year | The annual per capita rate at which individuals with SNTB recover from their TB infection/disease in the absence of treatment. | Truncated normal distribution, range: 0.073 - 0.209, SD: 0.029. | As preceding parameter |
| Relative risk of reinfection while latently infected (compared to de novo infection) | The risk of reinfection among individuals with latent TB infection, relative to those never previously infected. | Beta distribution [18] with α = 3.0, β = 8.0. | - The TB incidence rate was 13.5 per 1000 person-years (95% CI: 5.0–26.2) for reinfection and 60.1 per 1000 person-years (95% CI: 38.6–87.4) for primary infection [19].  To capture non-zero relative risk with a peak at 0.2 and a tail towards 1.0. |
| Relative risk of reinfection after recovery (compared to de novo infection) | The risk of reinfection among individuals who have recovered from active TB, relative to those never previously infected. | As for preceding parameter | As for preceding parameter |
| BCG vaccination coverage | Proportion of newborns receiving BCG vaccination each year. | Time-variant | WHO [20] |
| Reduced susceptibility to infection due to BCG vaccination | Reduction in the risk of TB infection among BCG-vaccinated infants and young children, with protection assumed to wane with age. | Age-specific | Assumed [4,21] |
| Rate of treatment commencement: shape of the hyperbolic tangent function | Defines the steepness of the logistic function, determining how gradually or abruptly the detection rate increases over time. | Uniform distribution, range: 0 - 0.5 | - Assumed.  - Minimally informative prior with bounds spanning the plausible range of values. |
| Rate of treatment commencement: Represents the point in time at which the detection rate undergoes the most rapid change, typically reflecting key historical interventions such as the establishment of the NTP and the introduction of DOTS. | Represents the point in time at which the detection rate undergoes the most rapid change, typically reflecting key historical interventions such as the establishment of the NTP and the introduction of DOTS. | Truncated normal distribution, range: 1986 – 2000, SD: 3.5. | - Assumed.  - To prevent sharp cut-offs at the boundaries while still capturing the plausible range of values |
| Rate of treatment commencement: Time from active TB to diagnosis and treatment commencement | The average delay from the onset of active TB disease to being diagnosed and starting treatment, influenced by diagnostic capacity and healthcare accessibility. | Truncated normal distribution, range: 0 - 10 with mean around 2 | - Assumed.  - To capture shorter delays while allowing for occasional longer delays  - To capture shorter delays while allowing for occasional longer delays |
| Average TB treatment duration | The average length of standard TB treatment, used to estimate outcomes such as treatment success, death, or relapse over time. | 0.5 years | Assumed [22] |
| Proportion of TB-related deaths during treatment | The proportion of TB deaths among individuals who do not successfully complete treatment. | 0.026 | [23] |

**2.2. Calibration**

To ensure our parameter starting distributions were widely dispersed across the multidimensional space, we first used Latin hypercube sampling [24] to generate 16 parameter set samples. The best eight samples, selected based on their log-likelihood, were then briefly optimized using a non-gradient-based method, run for 1000 iterations. These optimized parameter sets served as the basis for the main Bayesian sampling algorithm using the PyMC implementation of the differential evolution Metropolis algorithm “DEMetropolis(Z)” [25]. This algorithm enhances sampling efficiency by using randomly selected past samples from the Metropolis chains to inform new proposals, thereby improving the exploration of parameter space and ensuring more robust convergence. This proceeded for 50,000 tuning cycles followed by 100,000 sampling cycles. After the calibration phase, to obtain our final results with associated uncertainty ranges, we ran the model with 1,000 parameter sets from the accepted samples of the main calibration phase to generate uncertainty ranges and 95% credible intervals (CrIs, i.e. range from the 0.025^th^ to the 0.975^th^ quantile for each output). The best assumption concerning the impact of COVID-19 on TB dynamics was identified by comparing the expected log pointwise predictive density Watanabe-Akaike Information Criterion (EPLD-WAIC) values across four scenarios, with less negative values indicating better predictive performance.

Each candidate epidemiological model, consisting of 60 compartments (derived from 6 base compartments, stratified by 6 age groups and 3 clinical forms of TB), completed 8 chains with 150,000 iterations per chain within 4 hours on a computer with a 16-core 3rd Generation Intel Core i9-13900HK processor running at 3.5 GHz and 16 GB of RAM.

We evaluated the potential impact of future interventions to enhance case detection for TB between 2025 and 2035. The model simulated 2-fold, 5-fold increases in the rate at which individuals with active TB disease are diagnosed and initiated on treatment or simply the rate of commencing treatment. For each scenario, we estimated the total number of new TB cases and TB-related deaths averted.

**Table S3. Calibration Targets**

| **Targets** | **Values** | **Evidence** |
| --- | --- | --- |
| Total population | 2019: 96,484,000 | National census [26] |
| TB notifications | 2012: 103,906  2013: 102,196  2014: 102,087  2015: 100,780  2016: 102,097  2017: 102,725  2018: 99,658  2019: 102,503  2020: 99,582  2021: 77,657  2022: 102,479  2023: 104,517 | WHO [27] |
| Proportion of pulmonary among notifications | 2022: 83%  2023: 75% | WHO [27] |
| Prevalence of pulmonary TB among adults | 2017: 322 | The second national prevalence survey [28] |

### III. Additional results

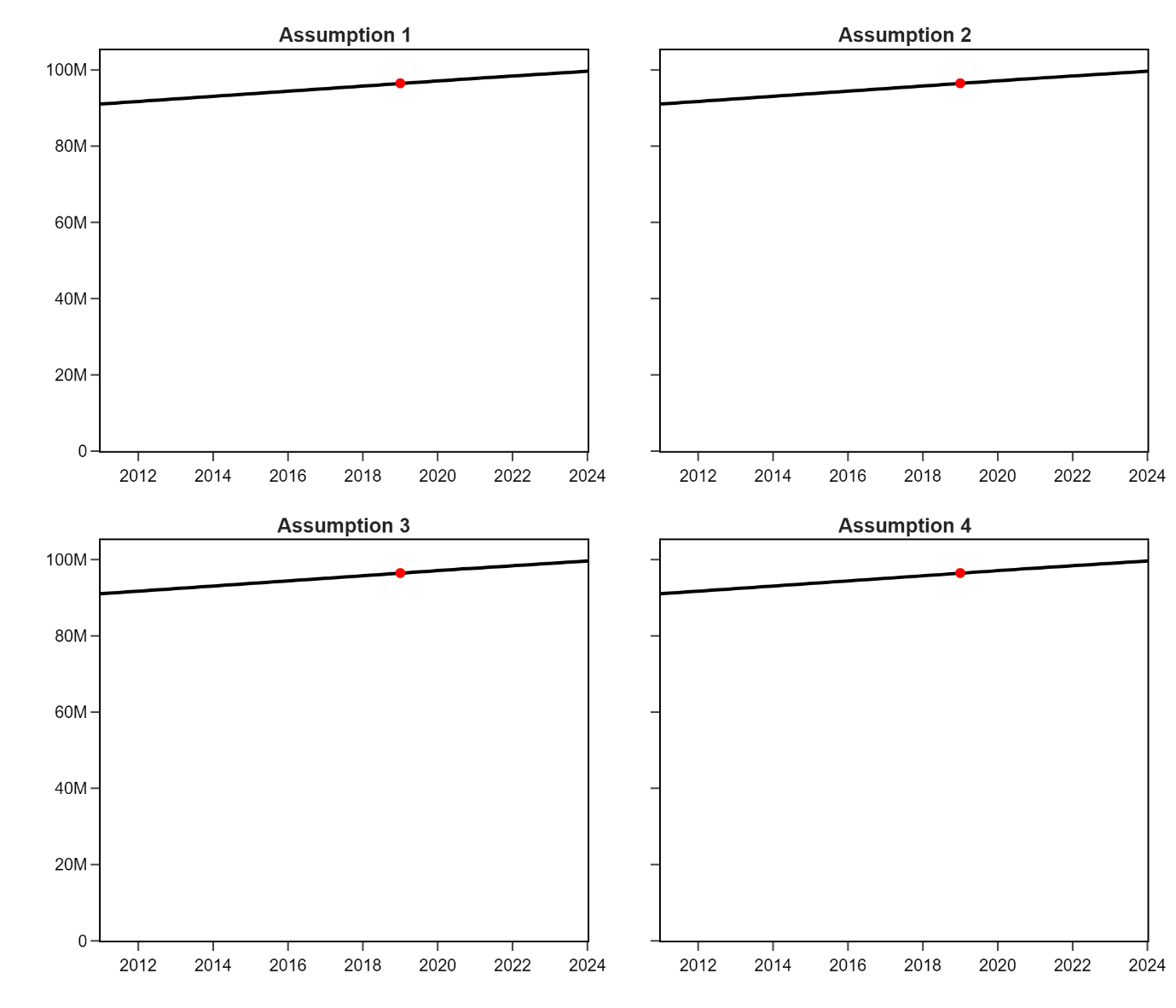

**Figure S3. Model fits to the total population under our four candidate assumptions regarding the effect of COVID-19 on *M.tb* dynamics.** *Assumption 1: The COVID-19 pandemic had no impact on TB case detection or transmission. Assumption 2: Only case detection was reduced. Assumption 3: Only TB transmission was reduced. Assumption 4: Both case detection and TB transmission were reduced, incorporating effects from both social distancing and healthcare disruption*.

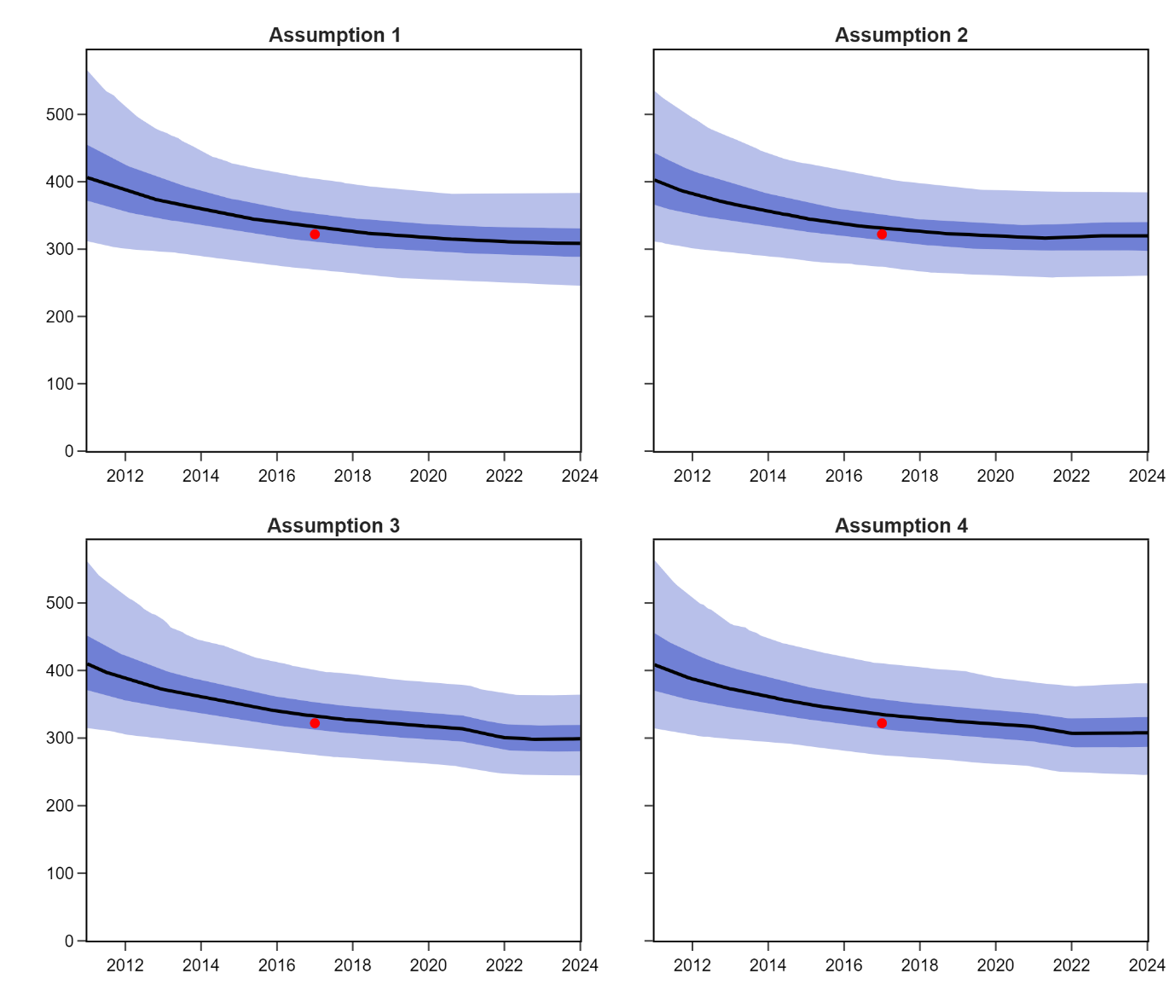

**Figure S4. Model fits to the prevalence of TB among adults under our four candidate assumptions regarding the effect of COVID-19 on *M.tb* dynamics.** *Assumption 1: The COVID-19 pandemic had no impact on TB case detection or transmission. Assumption 2: Only case detection was reduced. Assumption 3: Only TB transmission was reduced. Assumption 4: Both case detection and TB transmission were reduced, incorporating effects of both social distancing and healthcare disruption*.

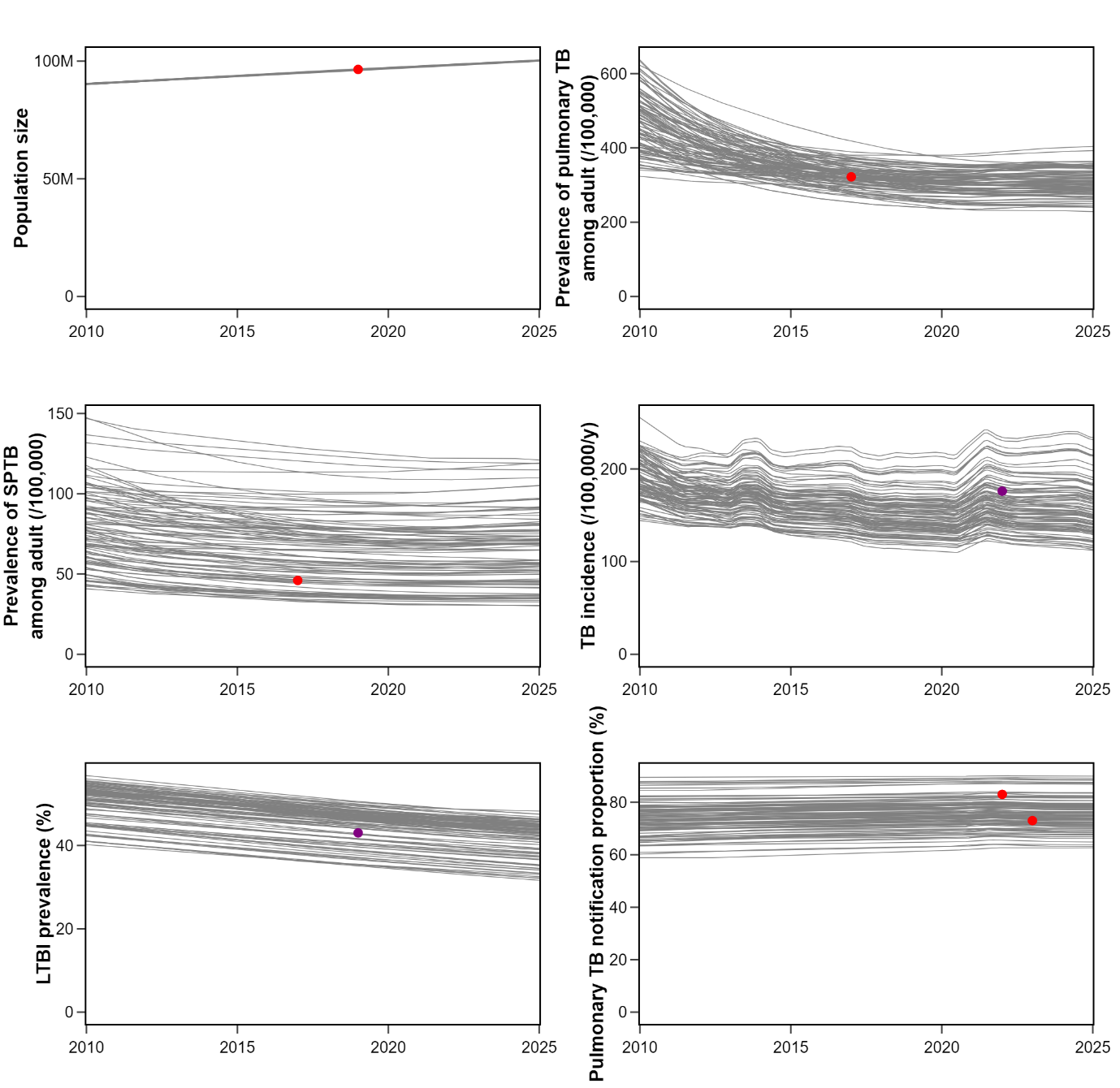

**Figure S5. Individual model trajectories generated from 100 randomly selected accepted parameter sets.** *Red points representing calibration targets and purple points indicating literature estimates excluded from calibration targets.*

**Table S4. Comparison of goodness-of-fit of 4 assumptions for COVID-19**

| **Assumption*** | **EPLD-WAIC** | **Expected number of parameters** |
| --- | --- | --- |
| Assumption 1 | -28.010 | 1.026 |
| Assumption 2 | -27.048 | 1.146 |
| Assumption 3 | -28.379 | 1.126 |
| Assumption 4 | -28.632 | 1.024 |

****Assumption 1:*** *The COVID-19 pandemic had no impact on TB case detection or transmission.* ***Assumption 2:*** *Only case detection was reduced.* ***Assumption 3:*** *Only TB transmission was reduced.* ***Assumption 4:*** *Both case detection and TB transmission were reduced, incorporating effects from both social distancing and healthcare disruption.* ***EPLD-WAIC:*** *expected log pointwise predictive density Watanabe-Akaike Information Criterion.*

**Table S5. Posterior estimates of calibrated parameters for the candidate with Assumption 2 (only case detection was reduced during COVID-19)**

| **Parameter** | **Mean** | **SD** | **ESS tail** | $\hat{\boldsymbol{R}}$ | **HDI** |
| --- | --- | --- | --- | --- | --- |
| Contact rate | 0.023 | 0.006 | 232.0 | 1.05 | 0.013 to 0.034 |
| Relative risk of infection for individuals with latent infection | 0.18 | 0.076 | 534.0 | 1.05 | 0.054 to 0.323 |
| Relative risk of infection for individuals with history of TB disease | 0.268 | 0.127 | 862.0 | 1.05 | 0.049 to 0.501 |
| Uncertainty multiplier for the rate of latent TB progression | 0.994 | 0.155 | 833.0 | 1.02 | 0.721 to 1.289 |
| Proportion of pulmonary TB among incidence | 0.761 | 0.057 | 681.0 | 1.03 | 0.654 to 0.872 |
| Proportion of SPTB among pulmonary incidence | 0.323 | 0.067 | 815.0 | 1.03 | 0.201 to 0.438 |
| SPTB death rate | 0.385 | 0.024 | 660.0 | 1.05 | 0.338 to 0.425 |
| SNTB TB death rate | 0.026 | 0.004 | 698.0 | 1.03 | 0.019 to 0.032 |
| SPTB self-recovery rate | 0.228 | 0.023 | 757.0 | 1.03 | 0.183 to 0.266 |
| SNTB TB self-recovery rate | 0.136 | 0.022 | 834.0 | 1.04 | 0.097 to 0.18 |
| Screening shape | 0.302 | 0.116 | 1165.0 | 1.03 | 0.109 to 0.5 |
| Screening inflection time | 1998.49 | 2.648 | 1008.0 | 1.02 | 1993.452 to 2003.141 |
| Time from active TB to be diagnosed | 1.812 | 0.463 | 1193.0 | 1.01 | 0.96 to 2.651 |
| Relative reduction of screening rate during COVID-19 | 0.303 | 0.18 | 664.0 | 1.05 | 0.01 to 0.608 |

*SD: Standard deviation, ESS: Effective sample size,* $\hat{R}$*: Gelman-Rubin statistics, HDI: High density interval*

*TB: Tuberculosis, SPTB: Smear-positive tuberculosis, SNTB: Smear-negative tuberculosis.*

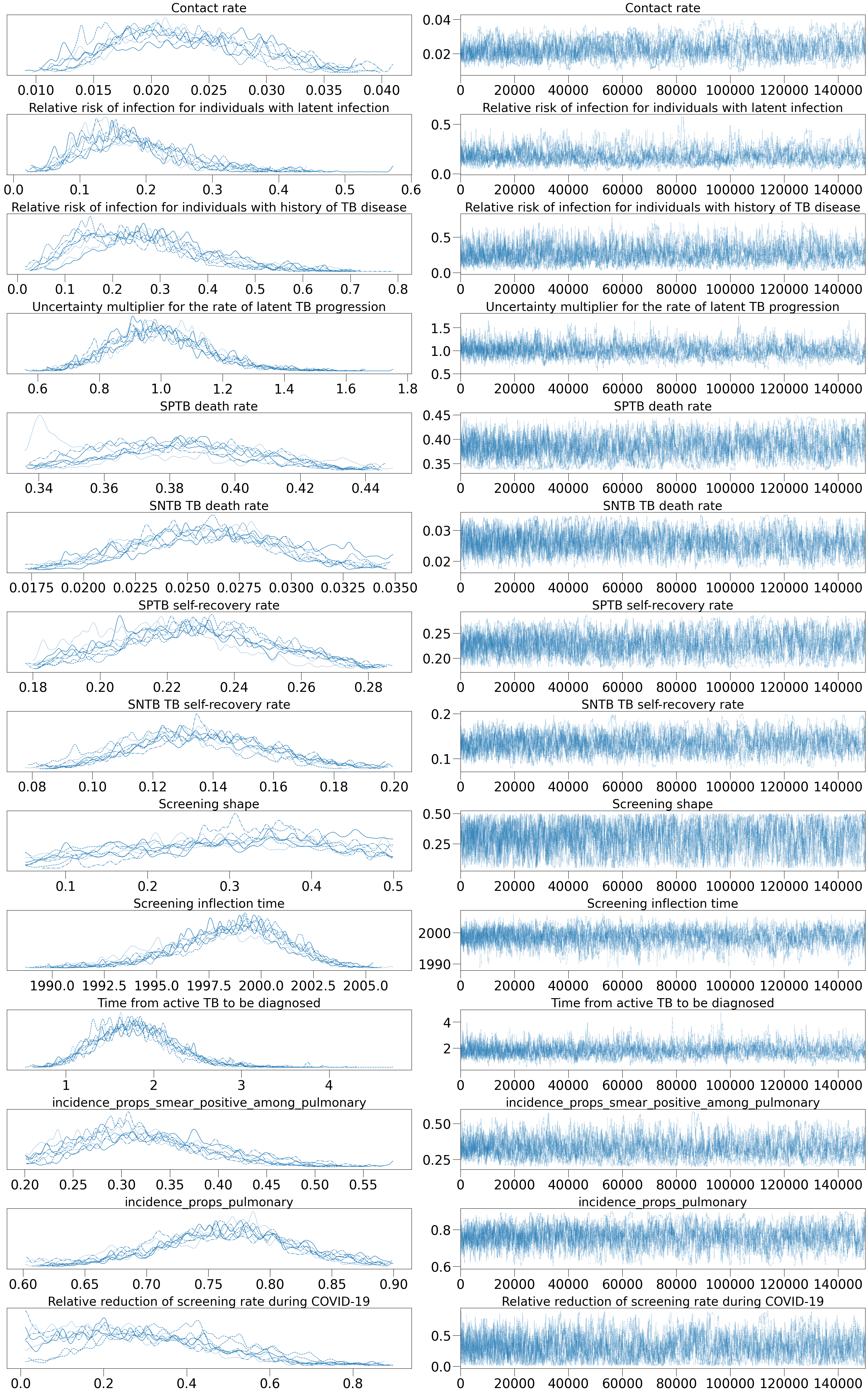

**Figure S6. Calibration parameter traces under Assumption 2.** *TB: Tuberculosis, SPTB: Smear-positive tuberculosis, SNTB: Smear-negative tuberculosis.*

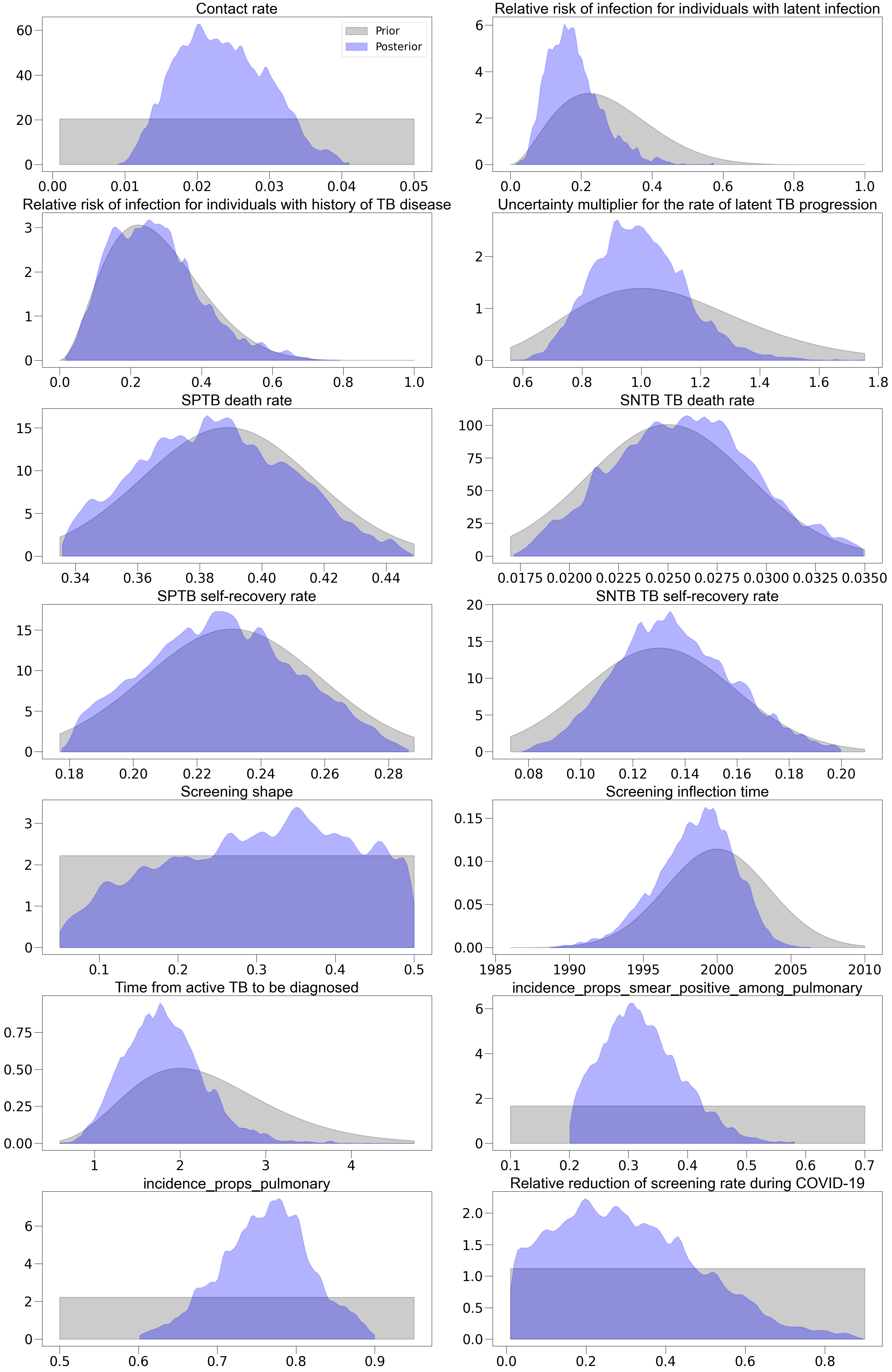

**Figure S7. Comparison of prior and posterior distributions of Assumption 2.** *TB: Tuberculosis, SPTB: Smear-positive tuberculosis, SNTB: Smear-negative tuberculosis.*

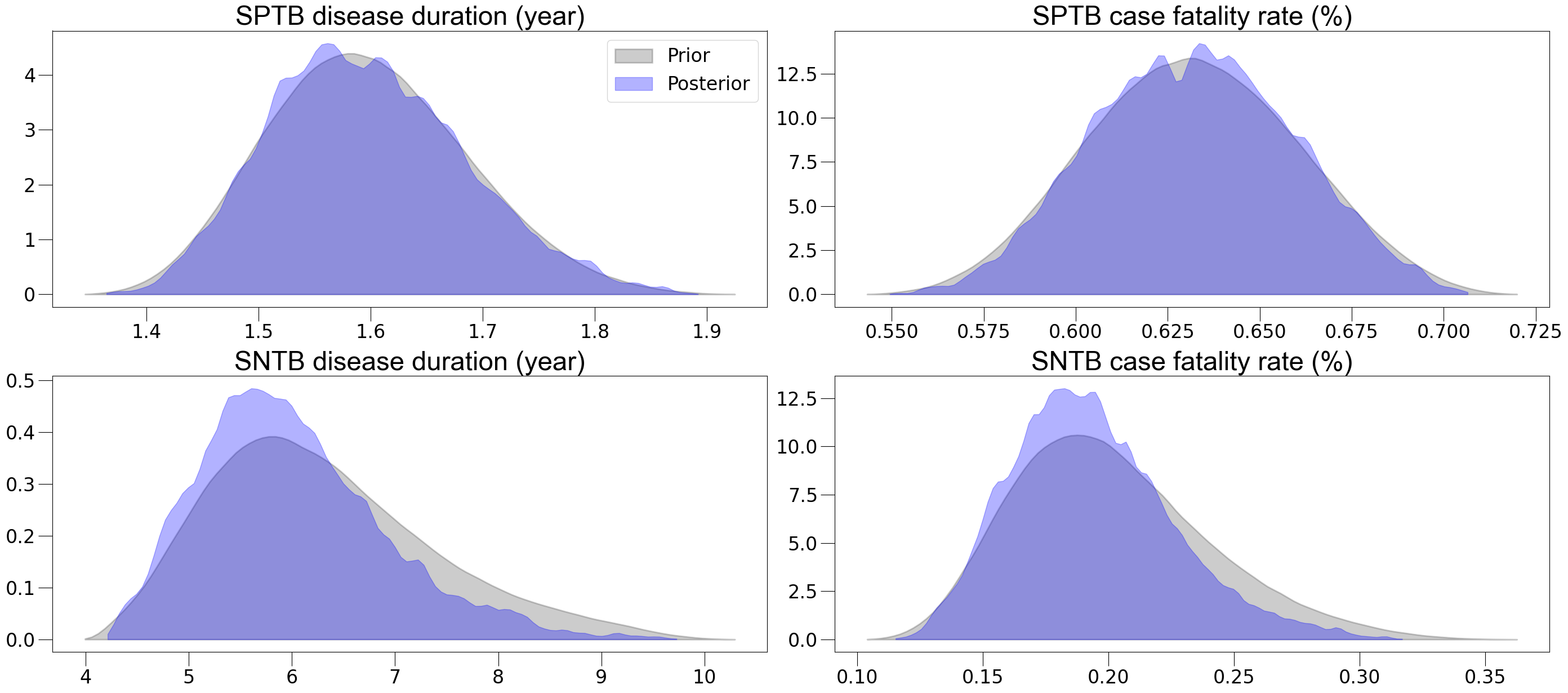

**Figure S8. Comparison of prior and posterior distributions for disease duration and case fatality rate without treatment of smear-positive and smear-negative TB.** *SPTB: smear-positive TB, SNTB: smear-negative TB.*

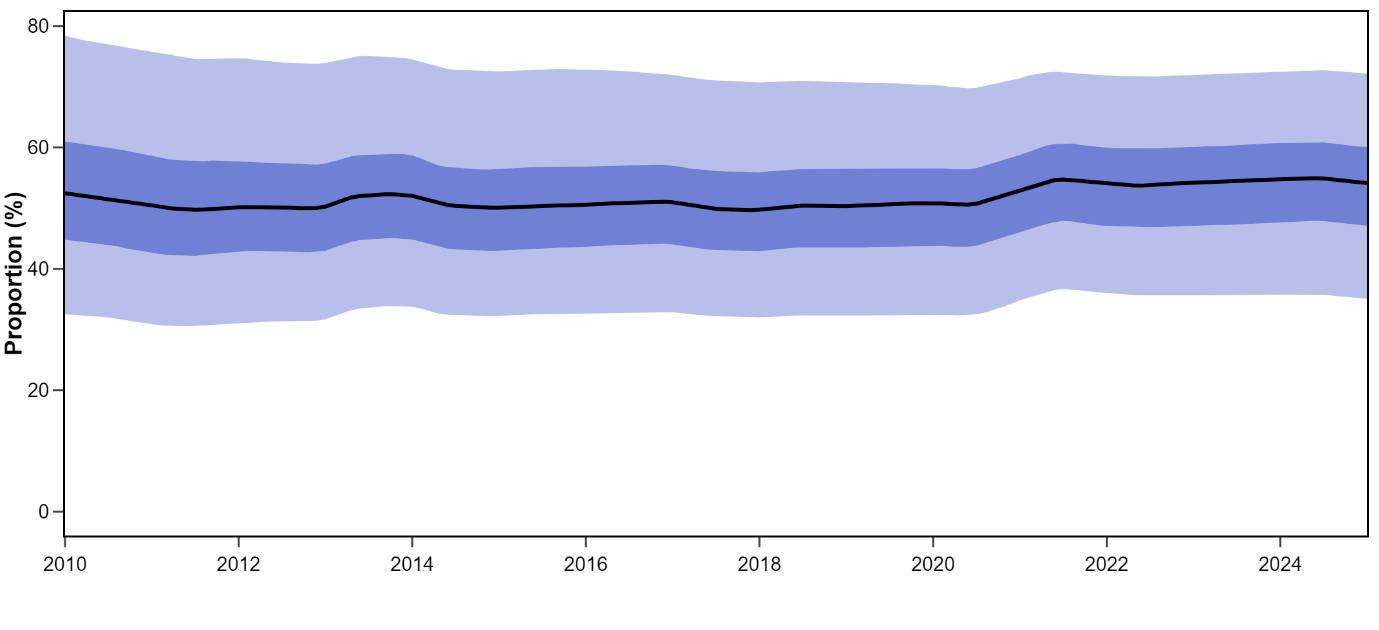

**Figure S9. Proportion of TB cases attributable to recent transmission.** *Solid lines represent median model estimates. Shaded areas show corresponding interquartile ranges (dark shade) and 95% credible intervals (light shade).*

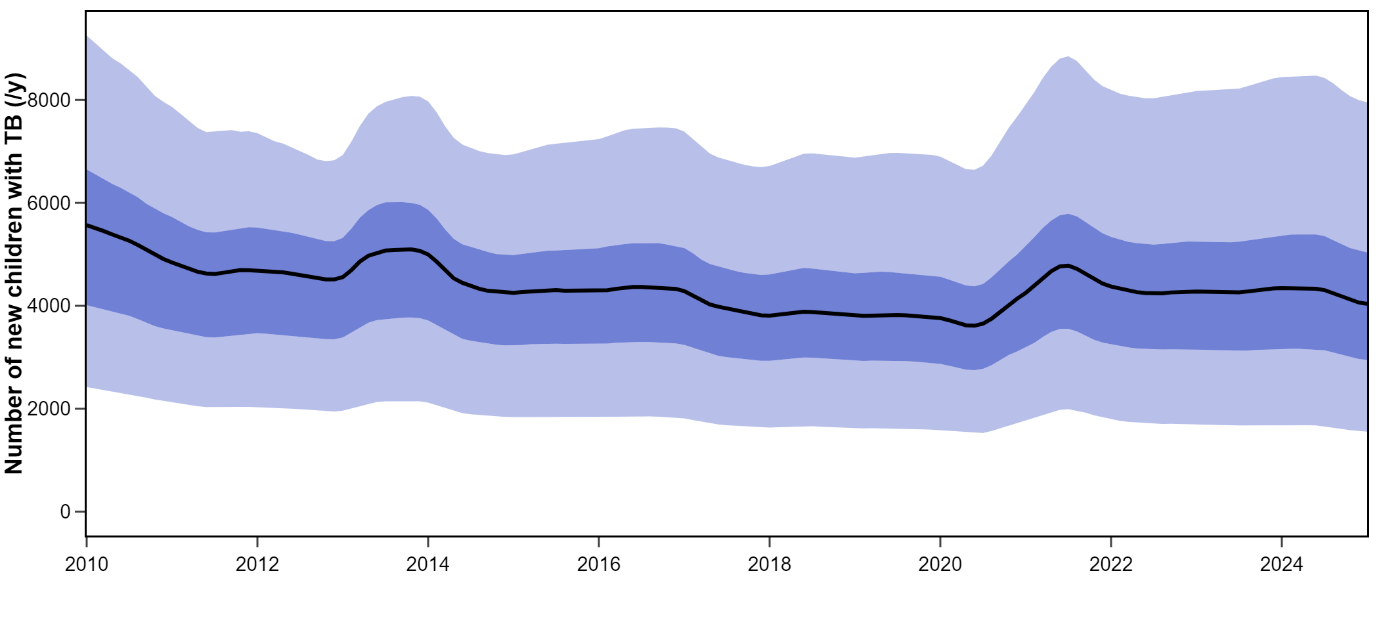

**Figure S10. Estimated number of new children with TB per year.** *Solid lines represent median model estimates. Shaded areas show corresponding interquartile ranges (dark shade) and 95% credible intervals (light shade).*

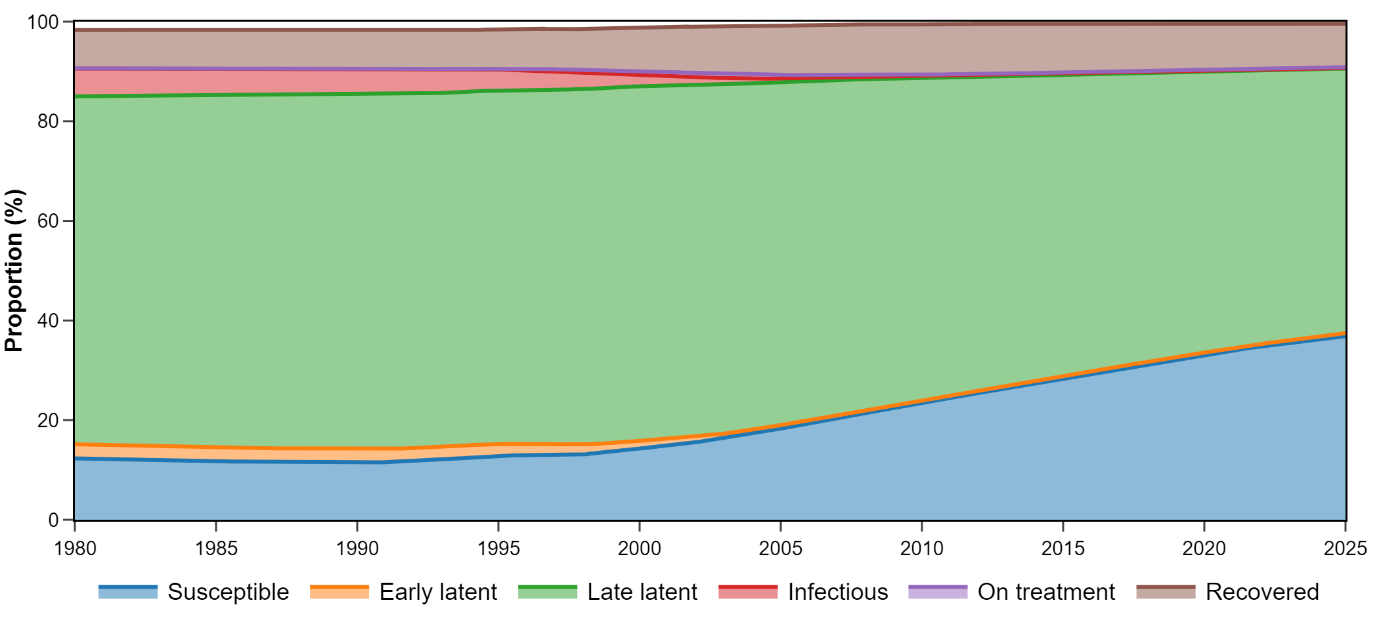
**Figure S11. Proportions of the population by compartment based on results from the maximum likelihood run.**

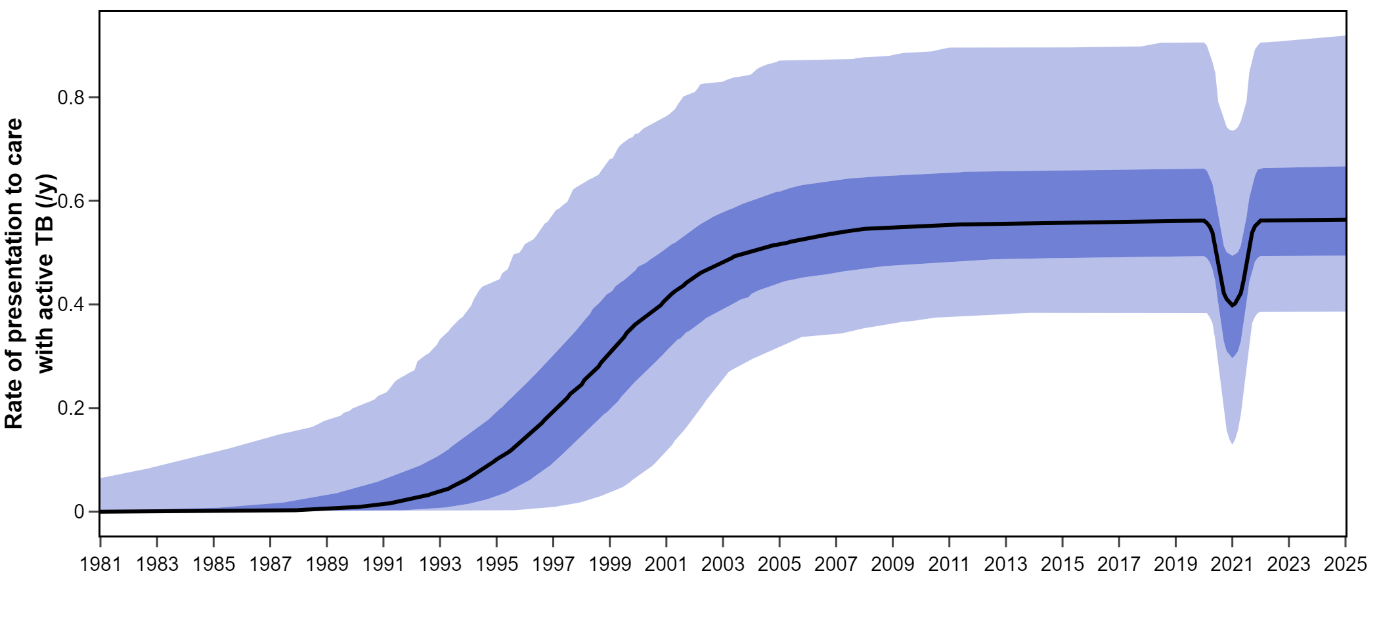

**Figure S12. Posterior modelled profile of the** **rate of commencing care of active TB.** Solid lines represent median estimates. Shaded areas show interquartile ranges (dark shade) and 95% credible intervals (light shade).

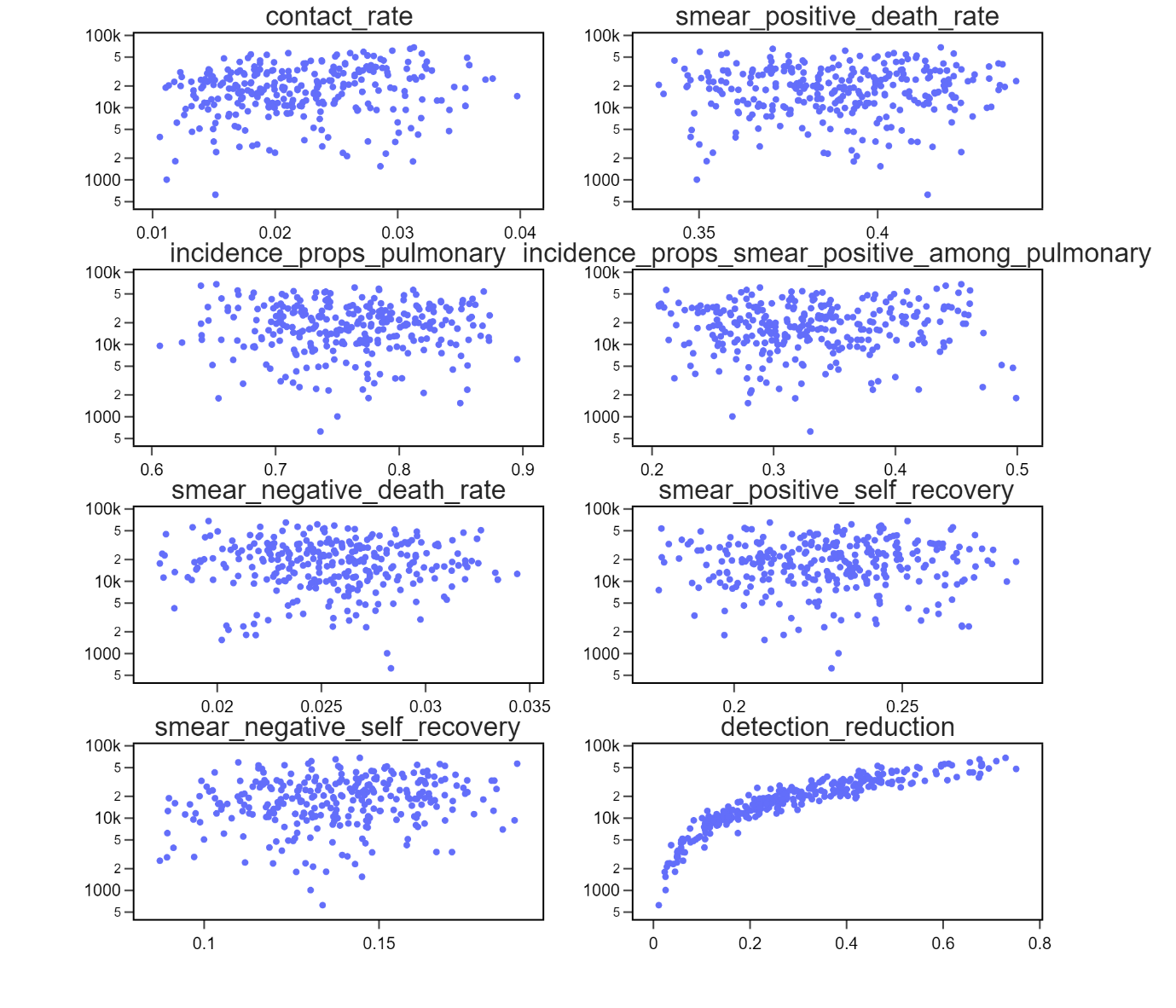

**Figure S13. Estimated cumulative number of new TB episodes plotted against key model parameters from 300 simulation runs.** *Values on y axes are presented in logarithmic scale.*

**REFERRENCES**

**Figure S1.**

**Alt text:** Figure S1 depicts an annual contact matrix segmented by age groups, showing the average number of interactions each year among various age categories. Each row corresponds to the age group of individuals reporting their contacts, while each column reflects the age groups of those they interacted with. The numbers in each cell of the matrix represent the total yearly interactions between individuals from the corresponding row and column age groups.

**Figure S2.**
**Alt text:** The presumed declining profile of the BCG vaccine's effectiveness over time is shown in Figure S2. The efficacy estimates from prior studies are shown by the black dashed line and the green dashed line, respectively. A visual comparison of various research findings on the effectiveness of the BCG vaccine is provided by the red line, which depicts the vaccine's modelled effect as it declines over time.

**Figure S3.**

**Alt text:** Model fits to the entire population under four potential assumptions on how COVID-19 affects the dynamics of *M.tb* are shown in Figure S3. Assumption 1, neither TB case detection nor transmission was impacted by the COVID-19 pandemic; Assumption 2, only case detection was decreased; Assumption 3, only TB transmission was decreased; and Assumption 4, decreases in both case detection and TB transmission.

**Figure S4.**

**Alt text:** Figure S4 depicts model fits to the prevalence of TB among adults under four different assumptions about the impact of COVID-19 on *M.tb* dynamics. Assumption 1, neither TB case detection nor transmission was impacted by the COVID-19 pandemic; Assumption 2, only case detection was decreased; Assumption 3, only TB transmission was decreased; and Assumption 4, decreases in both case detection and TB transmission.

**Figure S5.**

**Alt text:** Individual model trajectories produced from 50 approved parameter sets that were chosen at random are displayed in Figure S5. Every trajectory is shown as a line that illustrates how parameter uncertainty affects model outcomes. On the graph, purple points denote estimates from the literature that were left out of the calibration targets, while red points reflect calibration targets - specific data points that the model seeks to match. This graphic makes it easier to see how the model's outputs match important information from the literature and calibration.

**Figure S6**

**Alt text:** Calibration parameter traces are shown in Figure S6 under Assumption 2, which assumes that COVID-19 simply decreased the identification of TB cases. Under the given assumption, each trace line illustrates the variability and convergence of parameters unique to each form of tuberculosis by representing the parameter values investigated during the model calibration procedure.

**Figure S7.**

The prior and posterior distributions under Assumption 2, which postulates that COVID-19 predominantly affected TB case detection, are contrasted in Figure S7. The model fitting process highlights the changes in ideas about the parameters, and each set of distributions shows how the initial assumptions (priors) are updated to match the observed data (posteriors).

**Figure S8.**

**Alt text:** The prior and posterior distributions for the disease duration and case fatality rate without treatment for smear-positive and smear-negative tuberculosis are compared in Figure S8. The histograms show how the original model assumptions (priors) were modified to match the empirical data (posteriors).

**Figure S9.**

**Alt text:** Figure S9 shows a graph showing the proportion of TB cases associated with recent transmission. It highlights how early latent infections contribute to the total burden of active TB patients by giving a visual depiction of the dynamics of the transition from early latent to active TB.

**Figure S10.**

**Alt text:** Figure S10 shows the population's proportions allocated among the compartments, based on the outcomes of the model's maximum likelihood run. The chart visually segments the population into various health states or compartments as defined by the model, illustrating how each compartment is proportionally represented within the overall population.

**Figure S11.**

**Alt text:** Line plot showing the estimated annual number of new tuberculosis (TB) cases among children. Solid lines indicate the median values from model simulations. Dark shaded regions represent the interquartile range (25th to 75th percentile), while light shaded regions show the 95% credible intervals, reflecting the uncertainty around the estimates.

**Figure S12**

**Alt text:** Figure S12 displays the posterior modelled profile of the rate at which individuals commence care for active TB. The median estimates of this rate are shown on the graph as solid lines, and the uncertainty surrounding these estimates is shown as shaded areas. The 95% CrIs are represented by lighter shaded areas, which demonstrate the estimates' wider uncertainty range, while the darker shaded areas show the interquartile ranges, which give an indication of central tendency and dispersion.

**Figure S13.**

**Alt text:** Scatter plot showing the estimated cumulative number of new TB episodes from 300 simulation runs, plotted against key model parameters. Each line represents one run. The y-axis is displayed on a logarithmic scale to capture a wide range of outcome values.
